# Higher Physical Fitness Regulates *In Vitro* Tumor Cell Growth in Older Adults with Treatment Naïve Chronic Lymphocytic Leukemia (CLL)

**DOI:** 10.1101/2021.03.01.21252293

**Authors:** Andrea Sitlinger, Michael A. Deal, Erwin Garcia, Dana K. Thompson, Tiffany Stewart, Grace A. MacDonald, Nicolas Devos, David Corcoran, Janet S. Staats, Jennifer Enzor, Kent J. Weinhold, Danielle M. Brander, J. Brice Weinberg, David B. Bartlett

**Affiliations:** Division of Medical Oncology, Duke University School of Medicine, Durham, NC, USA; Duke Molecular Physiology Institute, Duke University School of Medicine, Durham, NC, USA; Hematologic Malignancies and Cellular Therapy, Duke University School of Medicine, Durham, NC, USA; Laboratory Corporation of America Holdings, Morrisville, NC, USA; Division of Hematology, Duke University School of Medicine and VA Medical Center, Durham, NC, USA; Duke Center for Genomic and Computational Biology, Duke University, Durham, NC, USA; Division of Surgical Sciences, Duke University School of Medicine, Durham, NC, USA

**Keywords:** Physical Fitness, NMR lipoprotein and inflammation, NGS exosomal miRNA, NK-cell phenotype

## Abstract

Chronic lymphocytic leukemia (CLL) is associated with physical dysfunction and low overall fitness that predicts poor survival following commencement of treatment. However, it remains unknown whether higher fitness in CLL patients provides anti-oncogenic effects. We identified ten fit (CLL-FIT) and ten less fit (CLL-UNFIT) treatment-naïve CLL patients from 144 CLL patients who completed a set of physical fitness and performance tests. Patient plasma was used to determine its effects on *in vitro* 5-day growth/viability of three B-cell cell lines (OSU-CLL, Daudi and Farage). Plasma exosomal miRNA profiles, circulating lipids, lipoproteins, inflammation levels, and immune cell phenotypes were also assessed. CLL-FIT was associated with fewer viable OSU-CLL cells at Day 1 (*p=*0.003), Day 4 (*p=*0.001) and Day 5 (*p=*0.009). No differences between groups were observed for Daudi and Farage cells. Of 455 distinct exosomal miRNAs identified, 32 miRNAs were significantly different between groups. Of these, 14 miRNAs had ≤-1 or ≥1 log2 fold differences. CLL-FIT patients had 5 exosomal miRNAs with lower expression and 9 miRNAs with higher expression. CLL-FIT patients had higher HDL cholesterol, lower inflammation, and lower levels of triglyceride components (all *p*<0.05). CLL-FIT patients had lower frequencies of low-differentiated NKG2+/CD158a/b^neg^ (*p=*0.015 and *p*=0.014) and higher frequencies of NKG2A^neg^/CD158b+ mature NK-cells (*p=*0.047). Absolute numbers of lymphocytes including CD19+/CD5+ CLL-cells were similar between groups (*p*=0.359). Higher physical fitness in CLL patients is associated with altered CLL-like cell line growth *in vitro*, and with altered circulating and cellular factors indicative of better immune functions and tumor control.

## Introduction

Chronic lymphocytic leukemia (CLL) is the most prevalent adult leukemia in the USA [1, 2], with a median age at diagnosis of approximately 70 years [3, 4]. Median overall survival is approximately 10 years, with durations ranging from months to decades [5]. The presentation of CLL is diverse, and patients have a shorter life expectancy than age-matched healthy populations [6]. CLL increases the risk of secondary malignancies and autoimmune diseases, and infections are the leading cause of death [7-9]. At present, there is no survival benefit from immediate or early therapy prior to established treatment indications, and most patients have a period of observation before therapy initiation [10, 11]. During the treatment naïve period, patients with CLL can have low overall fitness and physical dysfunction, both of which predict poor survival following commencement of treatment [12]. To-date, we are aware of no studies that have assessed the role physical fitness has on underlying pathophysiological factors of CLL [13].

Higher physical fitness and physical activity levels for lymphoma patients are associated with improvements in therapy-related side effects, physical functioning, and quality of life [13-15]. Although the underlying mechanisms are not fully understood, these effects can be partly explained by systemic changes or differences in host pathways including metabolism, inflammation and immune function that promote a less carcinogenic milieu [16]. In healthy adults, increasing physical fitness following diet and exercise training can result in reduced *in vitro* growth of breast and prostate cancer cell lines using autologous serum [17, 18]. In contrast, others reported that breast cancer survivors’ autologous serum did not alter *in vitro* cell line growth following a 6-month exercise program [19]. However, they found that after completion of a single acute session of exercise, autologous serum blunted *in vitro* growth of breast [19] and colorectal cancer cell lines [20]. In these studies, acute exercise was associated with a transient increase of IL-6, IL-8, and TNFα, but not for insulin, suggesting complex interactions of endocrine and inflammatory mediators on cell line growth *in vitro*. Since elevated levels of pro-inflammatory mediators and insulin pathways are hallmarks of certain cancers, including CLL [21-23], other physical fitness host pathways are also likely to be influencing cancer behavior *in vivo* [24].

The primary aim of our current study was to evaluate the role of high and low physical fitness plasma from treatment naïve CLL patients on the *in vitro* growth/survival of a CLL-like cell line (OSU-CLL) [25]. The secondary aim was to identify similarities and differences in (a) circulating factors and (b) mononuclear cell phenotypes associated with CLL pathology. We hypothesized that plasma from high physically fit CLL patients would reduce the growth/survival of OSU-CLL cells *in vitro*.

## Materials and Methods

### Patient Characteristics

Ten physically fit (CLL-FIT) and 10 least physically fit (CLL-UNFIT) treatment naïve CLL patients matched by age (mean ± SD: 67.8 ± 10.8 yrs., range: 51 – 86 yrs.) and sex (5M/5F in each group) were assessed for this study. Study participants were identified from 144 CLL patients assessed for physical fitness and function during their regular clinical visits at the Duke Cancer Center between July 2017 and March 2018. Of these patients, 69 were confirmed treatment naïve and Rai Stage 0-1. All participants gave written informed consent, and the study was approved by the Duke University Medical Center Institutional Review Board.

### Clinical Characteristics

Clinical indices were obtained from patient’s medical records. These included the CLL-IPI score, calculated as previously described [26], disease duration, cytogenetics (i.e. standard karyotype and fluorescent in situ hybridization for CLL), IGHV mutation status, and CD38 expression. Plasma levels of soluble CD20 (sCD20) and intercellular adhesion molecule 1 (ICAM-1) were determined in duplicate using a human sandwich immunoassay according to the manufacturer’s instructions (Meso Scale Discovery, Rockville, MD). β2-microglobulin (B2M) was determined in duplicate using a commercially available ELISA (R&D Systems, Minneapolis, MN). The lower limits of detection (LLOD) were sCD20 (30.5 pg/mL), ICAM-1 (2.60 ng/mL), and B2M (0.132 µg/mL). All samples had concentrations greater than the LLOD with the exception of sCD20 with 81% of samples above the LLOD. Complete blood counts (CBCs) and differentials were clinically assessed at the Duke Clinical Laboratory using an automated hematology analyzer (Sysmex, Lincolnshire, IL, USA).

### Physical Performance and Fitness

Patients completed a short battery of standardized physical performance tests. These included the 6-minute walk test (6MWT), the short physical performance battery (SPPB), timed-up and go (TUG), and grip strength.Following completion of physical testing, patients completed two physical activity questionnaires – the Incidental and Planned Activity Questionnaire (IPAQ) and the Stanford Brief Activity Survey (SBAS). We measured height and weight before testing, and blood pressure and resting heart rate following 10 minutes of seated rest. The eVO_2peak_ (mL/kg/min) was calculated using a validated equation that incorporates 6MWT distance, resting heart rate, weight, sex, and age [27]. We stratified groups using eVO_2peak_ matched for age (±3 yrs.) and gender.

### Blood Sampling

Approximately 20 mL of blood was collected into vacutainers containing either EDTA or heparin as an anti-coagulant. Blood was centrifuged at 3000 *x rpm* for 10 minutes at 4°C, and 3-4 mL of plasma, aliquoted, and immediately frozen at −80°C. From the remaining blood, mononuclear cells (PBMCs) were isolated using Ficoll-Hypaque (GE Life Sciences, PA) density centrifugation, and stored in aliquots of 10 × 10^6^ cells/mL in 90% fetal bovine serum (FBS) plus 10% DMSO, in the vapor phase of liquid nitrogen [28].

### Autologous Plasma Incubation with Cell Lines

We acquired the OSU-CLL cell line, generated by EBV transformation, from the Byrd Lab at Ohio State University [25]. This cell line is characterized by CD5 positivity, mutated IGHV, trisomy 12 and trisomy 19, a non-complex karyotype and wild type p53 expression. Importantly, it is stable under extended periods of culture. In addition to the OSU-CLL line, we used two other B-cell lymphoma cell lines to assess potential similarities between cell growth and plasma characteristics. Specifically we used the Daudi (ATCC^®^ CCL-213) and Farage (ATCC^®^ CRL-2630) cell lines. The Daudi line is an EBV transformed Burkitt’s lymphoma B-lymphocyte, while the Farage is an EBV transformed mature B-cell iine from a patient with diffuse large cell non-Hodgkin’s lymphoma. The Daudi line is CD19+, CD5^neg^, β2-microglobulin negative, and the Farage line is CD19+, CD20+, CD5^neg^, HLA-DR+. Cells were thawed and grown for 7 days in complete medium (RPMI + 56 U/mL penicillin + 56 µg/ml streptomycin + 2 mM L-glutamine) + 10% fetal bovine serum (FBS). Cells were then washed and resuspended at 5 × 10^5^ cells/mL in complete media + 10% FBS (Control), or had the FBS replaced with 10% autologous plasma. To determine whether the differences between controls (serum) and autologous plasma might be affected by anticoagulant agents, we also incubated serum (collected at the same time for a different study) from four patients with the OSU-CLL line. Correlations between autologous serum and plasma were r=0.85 (data not shown). At baseline and every 24 hours for 5 days, cell density was measured in duplicate by both hemocytometer counting and the cell-counting feature of the Attune NxT flow cytometer (ThermoFisher). Live viable cells were quantified as Annexin^neg^/PI^neg^, early apoptotic as Annexin^pos^/PI^neg^, and late apoptotic/necrotic as Annexin^pos^/PI^pos^ using manufacturer guidelines (BD Biosciences) using the Attune NxT flow cytometer.

### Exosomal miRNA

Plasma (500 µL) was briefly thawed at 37°C and filtered to exclude particles >0.8 µm for isolation of exosomal RNA using the Qiagen exoRNeasy midi kit (Qiagen, Hilden, Germany). We then purified exosomes before RNA extraction was completed using Qiagen miRNeasy chloroform based technology. RNA quantity and quality was quantified on a NanoDrop 2000 spectrophotometer (ThermoFisher). For sequencing, 5 uL of each exosomal RNA extract was used to generate a miRNA library using principals and methods similar to previously described [29-31]. The libraries were generated using the QIAseq miRNA Library kit (Qiagen). This kit uses modified adapters that efficiently ligate to microRNAs generated by Dicer processing. It also uses 12 bp Unique Molecular Indices (UMIs) to tag each miRNA at an early stage, eliminating PCR and sequencing bias. During library prep, libraries were indexed using a single 6 bp indexing approach allowing for multiple libraries to be pooled and sequenced on the same sequencing flow cell of an Illumina sequencing platform. Before pooling and sequencing, we assessed fragment length distribution and overall library quality on an Agilent Fragment Analyzer instrument (Agilent Technologies). Concentrations of each library were assessed using a Qubit fluorometer (ThermoFisher). We then pooled 20 libraries in an equimolar ratio and sequenced on a NextSeq High Output flow cell. To sequence the 12 bp UMIs that were used to tag each miRNA, sequencing was completed at a 75 bp single read. Approximately sixteen million reads were generated for each sample, and a total of 455 distinct miRNAs identified.

### miRNA Analyses

smRNA-seq data was processed using the TrimGalore toolkit [32] that employs Cutadapt [33] to trim low quality bases and Illumina sequencing adapters from the 3’ end of the reads. Only reads that were 18 to 28 nt in length after trimming were kept for further analysis. Reads were mapped to the hg19 version of the human genome using the Bowtie alignment tool [34]. Reads were kept for subsequent analysis if they map to no more than 13 genomic locations. Gene counts were compiled using custom scripts that compare mapped read coordinates to the miRBase [35] microRNA database. Reads that match the coordinates of the known mature microRNAs were kept if they perfectly matched the coordinates of the miRNA seed while not varying by more than 2 nt on the 3’ end of the mature miRNA. Only mature miRNAs that had at least 10 reads in any given sample were used in subsequent analysis. Normalization and differential expression were carried out using the DESeq2 [36] Bioconductor [37] package from the *R* statistical programming environment. Normalization was performed using the ‘*poscounts*’ approach to eliminate systematic differences across the samples.

### Flow Cytometry

PBMCs were partly thawed in a 37°C water bath, then completely thawed using the dropwise method and washed in 37°C thaw buffer (RPMI + 10% FBS + 1% penn/strep + 1% L-Glutamine + 25U/µL Benzonase). Cells were counted and resuspended at 10 × 10^6^ cells/mL in Dulbecco’s PBS (DPBS), and 100 µL aliquoted into FACS tubes. Cells were first stained with 0.1 µL of the viability dye Zombie Aqua (BioLegend, CA), before blocking Fc receptors (Human TruStain FcX, BioLegend), followed by complete combinations of the following antibodies. NK-cell tubes contained 0.63 µg/mL CD3 BUV395 (Clone SK7; BD Bioscience), 0.16 µg/mL CD56 BB700 (Clone NCAM16.2; BD Bioscience), 0.25 µL NKG2A PE (Clone REA110; Miltyeni Biotec, MD), 2.5 µL NKG2C PE-Vio770 (Clone REA205; Miltyeni Biotec), 10 µg/mL NKG2D BB515 (Clone 1D11; BD Bioscience), 5 µg/mL CD244 BV421 (Clone 2-69; BD Bioscience), 5 µL CD158a APC-Vio770 (Clone REA284; Miltyeni Biotec), and 5 µg/mL CD158b APC (Clone DX27; BioLegend). Monocyte tubes contained 1.25 µg/mL CD14 BUV805 (Clone M5E2; BD Bioscience) and 1.25 µg/mL CD16 BUV395 (Clone 3G8; BD Bioscience). T-cell and B-cell tubes contained 1 µg/mL CD3 Pacific Blue (Clone UCHT1; BD Bioscience), 1 µg/mL CD4 PE (Clone OKT4; BioLegend), 10 µg/mL CD8 FITC (Clone OKT8; Thermofisher, MA), 1.5 µg/mL CD19 APC-Cy7 (Clone HIB19; BioLegend), and 3 µg/mL CD5 APC (Clone UCHT2; BioLegend). We titrated all antibodies prior to assessing samples, and used single color and flow minus one (FMOs) tubes for compensation. Cells were incubated for 30 minutes on ice in the dark before being fixed with 1% paraformaldehyde (Sigma Aldrich). Cells analyzed on either a BD LSR Fortessa (NK-cell and monocytes) equipped with 4 lasers or a BD FACS Canto II (T-cell and B-cells) equipped with 3 lasers. All analyses were completed after acquisition using FCS Express v6 (DeNovo Software, CA).

### Nuclear Magnetic Resonance (NMR) Spectroscopy

Plasma (600 µL) stored at −80°C was analyzed by NMR at LabCorp (Morrisville, NC, USA) as single batch. NMR spectra were acquired on a Vantera® Clinical Analyzer as previously described [38]. The concentration of GlycA, a marker of systemic inflammation [39, 40], was calculated from NMR signal amplitudes of highly mobile protons of *N*-acetylglucosamine residues located on the carbohydrate side-chains of circulating acute phase proteins (e.g., α1-acid glycoprotein, haptoglobin, α1-antitrypsin, α1-antichymotrypsin, and transferrin). Concentrations of lipids, lipoprotein particles, apolipoproteins, and particle sizes were measured using advanced proprietary deconvolution algorithm (LP4) which provides better resolution of subclasses compared to previous algorithms [38, 41, 42]. We calculated the Lipoprotein Insulin Resistance Index (LP-IR) from NMR-measured lipoprotein particle sizes (TRL, LDL and HDL) and particle concentrations (very large TRL + large TRL, small LDL, large HDL). LP-IR scores range from 0 (least insulin resistant) to 100 (most insulin resistant) [38, 41]. Valine, leucine and isoleucine (BCAAs) and their sum (total BCAA) were quantified as previously described [43]. Glucose, glycine and alanine were measured using the LP4 algorithm.

### Statistical Analyses

We conducted patient characteristics, NMR data, and immune data analyses using SPSS version 23.0 (IBM, Armonk, NY, USA). Normality was assessed using Kolmogorov-Smirnov analysis. For variables violating normality, we used non-parametric analyses. Comparisons of variables were completed using Independent T-tests, Mann-Whitney U tests, and Chi-square analyses were used for categorical variables. Spearman correlations were conducted between variables as measures of associations. For analyses of cell line incubations with autologous plasma, a repeated linear mixed model was used to model both the numeric and percentage changes in cell growth. The model included main effects and interaction effects for time (Days 0, 1, 2, 3, 4, and 5) and group (CLL-FIT and CLL-UNFIT). Contrasts were used *a priori* to determine overall effects for time within each group, where significant effects for time occurred, and differences between groups at each time point. Results are presented with standard deviations (SD), 95% confidence intervals (CI) and effect sizes calculated as Cohen’s D (*d*). Statistical significance was accepted as p ≤ 0.05.

## Results

### Group Demographics, Clinical Measures, Physical Fitness and Function

Groups were clinically similar for staging, disease duration, and cytogenetics (all *p>*0.1) (Table 1). CLL-UNFIT patients were heavier (*p=*0.049), and had a higher BMI (*p=*0.013). CLL-FIT had higher cardiorespiratory fitness based on eVO_2peak_ (*p<*0.001), which was characterized by CLL-FIT completing 20% greater 6MWT distance (*p=*0.027) and 18% better on the SPPB (*p=*0.012). Both groups had similar grip strengths, TUG times, and similar self-reported exposure to physical activity levels (all *p>*0.05). CLL-UNFIT had higher neutrophil counts (*p=*0.046), while lymphocyte (*p=*0.517) and monocyte counts (*p=*0.694) were similar in both groups. No differences were observed between groups for absolute numbers of CD19+/CD5+ CLL B-cells (*p*=0.314), CD4+ (p=0.169) or CD8+ (*p*=0.438) T-cells. Similarly, no differences were observed for CD14+/CD16^neg^ (*p*=0.861), CD14+/CD16+ (*p*=0.635), or CD14+/CD16++ (*p*=0.598) monocytes (data not shown).

**Table 1:** Demographics and Clinical Characteristics

|  | CLL-FIT<br>(N=10) |  |  | CLL-UNFIT<br>(N=10) |  |  | p-value | 95% C.I. | Effect<br>Size (*d*) |
| --- | --- | --- | --- | --- | --- | --- | --- | --- | --- |
| **Demographics** |  |  |  |  |  |  |  |  |  |
| Age (years) | 65.8 | ± | 11.1 | 69.8 | ± | 10.8 | 0.426 | -14.3, 6.3 | 0.37 |
| Sex (M/F)<sup>1</sup> | 5/5 |  |  | 5/5 |  |  | 1.000 |  | 0.00 |
| Height (cm) | 167.8 | ± | 17.0 | 170.8 | ± | 11.9 | 0.648 | -16.8, 10.7 | 0.20 |
| Weight (kg) | 66.2 | ± | 16.8 | 85.8 | ± | 24.1 | **0.049** | -39.1, -0.1 | 0.94 |
| BMI (kg/m<sup>2</sup>) | 23 | ± | 2.9 | 29 | ± | 6.2 | **0.013** | -10.5, -1.4 | 1.24 |
| Resting Heart Rate (bpm) | 68 | ± | 12 | 72 | ± | 12 | 0.438 | -15.3, 6.9 | 0.33 |
| **Clinical Characteristics** |  |  |  |  |  |  |  |  |  |
| CLL-IPI Score | 2.4 | ± | 2.2 | 1.3 | ± | 1.1 | 0.175 | -.53, 2.74 | 0.63 |
| RAI Stage [N (%)]<sup>1</sup> |  |  |  |  |  |  |  |  |  |
| Stage 0 | 9 (90) |  |  | 7 (70) |  |  | 0.453 |  | 0.346 |
| Stage I | 1 (10) |  |  | 2 (20) |  |  |  |  |  |
| Unknown | 0 |  |  | 1 (10) |  |  |  |  |  |
| Disease Duration (years) | 5.0 | ± | 2.9 | 4.9 | ± | 2.7 | 0.937 | -2.5, 2.7 | 0.04 |
| Cytogenetics [N (%)]<sup>1</sup> |  |  |  |  |  |  |  |  |  |
| 13q Deletion | 7 (70) |  |  | 5 (50) |  |  | 0.361 |  | 0.20 |
| 17p Deletion | 3 (30) |  |  | 0 |  |  | 0.060 |  | 0.42 |
| 11q Deletion | 2 (20) |  |  | 0 |  |  | 0.136 |  | 0.33 |
| Trisomy 12 | 0 |  |  | 2 (20) |  |  | 0.136 |  | 0.33 |
| TP53 Mutated | 4 (40) |  |  | 1 (10) |  |  | 0.291 |  | 0.35 |
| IGHV Mutated<sup>1</sup> | 6 (60) |  |  | 6 (60) |  |  | 0.766 |  | 0.16 |
| CD38 Expression >30%<sup>1</sup> | 1 (10) |  |  | 0 |  |  | 0.119 |  | 0.46 |
| WBC Counts (x10<sup>3</sup>/μL) | 65 | ± | 54 | 51 | ± | 51 | 0.553 | -35.1, 63.5 | 0.27 |
| Lymphocytes | 59 | ± | 52 | 44 | ± | 49 | 0.517 | -32.7, 62.8 | 0.30 |
| CD19+/CD5+ CLL-cells | 48.1 | ± | 48.6 | 40.8 | ± | 41.2 | 0.359 | -35.0, 49.7 | 0.16 |
| CD4+ T-cells | 1.6 | ± | 2.3 | 5.1 | ± | 7.4 | 0.169 | -8.6, 1.6 | 0.64 |
| CD8+ T-cells | 0.8 | ± | 0.9 | 2.2 | ± | 2.3 | 0.088 | -3.0, 0.2 | 0.80 |
| Monocytes | 1.4 | ± | 1.4 | 1.8 | ± | 2.3 | 0.694 | -2.1, 1.5 | 0.21 |
| Neutrophils | 3.4 | ± | 1.6 | 4.9 | ± | 1.6 | **0.046** | -3.0, -0.3 | 0.94 |
| Neutrophil: T-cell | 3.7 | ± | 5.7 | 2.4 | ± | 2.5 | 0.510 | -2.2, 0.8 | 0.30 |
| T-cell: Monocyte | 2.1 | ± | 1.4 | 6.5 | ± | 7.6 | 0.086 | -9.6, 0.7 | 0.81 |
| Platelets (x10<sup>3</sup>/μL) | 149 | ± | 43 | 196 | ± | 62 | 0.065 | -96.4, 3.2 | 0.88 |
| Hemoglobin (g/dL) | 13.0 | ± | 1.7 | 13.6 | ± | 1.5 | 0.433 | -2.1, 0.92 | 0.37 |
| β2-microglobulin (mg/dL) | 1.9 | ± | 0.7 | 2.4 | ± | 0.9 | 0.228 | -1.3, 0.33 | 0.57 |
| ICAM-1 (ng/mL) | 709.1 | ± | 171.0 | 685.1 | ± | 158.6 | 0.755 | -136, 184 | 0.15 |
| sCD20 (pg/mL) | 220.8 | ± | 306.9 | 118.2 | ± | 181.3 | 0.417 | -158, 364 | 0.40 |
| **Fitness and Performance** |  |  |  |  |  |  |  |  |  |
| eVO<sub>2peak</sub> (mL/kg/min) | 34.2 | ± | 3.3 | 24.9 | ± | 3.2 | **<0.001** | 6.2, 12.4 | 2.86 |
| 6MWT (m) | 500 | ± | 77 | 399 | ± | 107 | **0.027** | 12.6, 188 | 1.08 |
| SPPB Score | 12 | ± | 0 | 9.9 | ± | 2.4 | **0.012** | .52, 3.68 | 1.24 |
| TUG (sec) | 9.4 | ± | 1.6 | 11.6 | ± | 4.5 | 0.170 | -5.6, 1.07 | 0.65 |
| Grip Strength Right (kg) | 33.3 | ± | 12.0 | 32.7 | ± | 16.7 | 0.930 | -13.1, 14.3 | 0.04 |
| Grip Strength Left (kg) | 32.4 | ± | 14.3 | 30.5 | ± | 16.1 | 0.780 | -12.4, 16.3 | 0.12 |
| Best Grip (BMI normalized) | 1.4 | ± | 0.4 | 1.1 | ± | 0.5 | 0.171 | -.15, .77 | 0.66 |
| IPAQ (total hours/week) | 31.9 | ± | 29.1 | 26.4 | ± | 16.8 | 0.610 | -10.2, 50.6 | 0.23 |
| SBAS [N (%)]<sup>1</sup> |  |  |  |  |  |  |  |  |  |
| Inactive | 2 (20) |  |  | 3 (30) |  |  | 0.936 |  | 0.21 |
| Light | 4 (40) |  |  | 3 (30) |  |  |  |  |  |
| Moderate | 2 (20) |  |  | 2 (20) |  |  |  |  |  |
| Hard | 1 (10) |  |  | 1 (10) |  |  |  |  |  |
| Very Hard | 1 (10) |  |  | 1 (10) |  |  |  |  |  |

<sup>1</sup>Chi-square Mean ± SD. BMI (Body Mass Index); CLL-IPI (CLL- International Prognostic Index); IGHV (Immunoglobulin Heavy Chain Gene); WBC (White Blood Cell); ICAM-1 (Intercellular Adhesion Molecule 1); sCD20 (soluble CD20); 6MWD (6-Minute Walk Test); SPPB (Short Physical Performance Battery); TUG (Timed-Up And Go); IPAQ (Incidental and Planned Activity Questionnaire); SBAS (Stanford Brief Activity Survey). Continuous variables are mean ± SD, nominal data is N (%)

### Cell Line Growth with Autologous Plasma

Cell growth was quantified by both the average cell count per well from duplicate wells and the percentage change of cell counts over time in culture (Figure 1). Results are representative from ten CLL-FIT and nine CLL-UNFIT patients. There was a significant main effect for time (F (5, 85) = 615.0; *p<*0.001; η^2^ =.973) and a group × time interaction (F (5, 85) = 5.17; *p<*0.001; η^2^ =.233) for viable OSU-CLL cell numbers (Fig. 1A). At Day 1 (*p=*0.003), Day 4 (*p=*0.001) and Day 5 (*p=*0.009) CLL-UNFIT had 16%, 12%, and 15.5% more cells than CLL-FIT did, respectively. Similarly, there was a significant main effect for time (F (4, 68) = 88.1; *p<*0.001; η^2^ =.838) and a group × time interaction (F (4, 68) = 6.57; *p<*0.001; η^2^ =.279) for viable OSU-CLL percentage change from the previous day (Fig. 3B). Compared to CLL-FIT, at Day 1 (*p=*0.010) and Day 4 (*p=*0.016) CLL-UNFIT had a 42.5% and 7.7% greater increase in previous day cell numbers, but a 16.5% lower increase in previous cell numbers at Day 2 (*p=*0.042). There was a significant main effect for time for Daudi cell numbers (Fig. 3C: F (5, 85) = 770.0; *p<*0.001; η^2^ =.980) and percentage change (Fig. 3D: F (5, 85) = 192.2; *p<*0.001; η^2^ =.923) but no group × time interactions. Similarly, there was a significant main effect for time for Farage cell numbers (Fig. 3E: F (5, 85) = 165.9; *p<*0.001; η^2^ =.907) and percentage change (Fig. 3F: F (5, 85) = 16.2; *p<*0.001; η^2^ =.489) but no group × time interactions.

**Figure 1.**
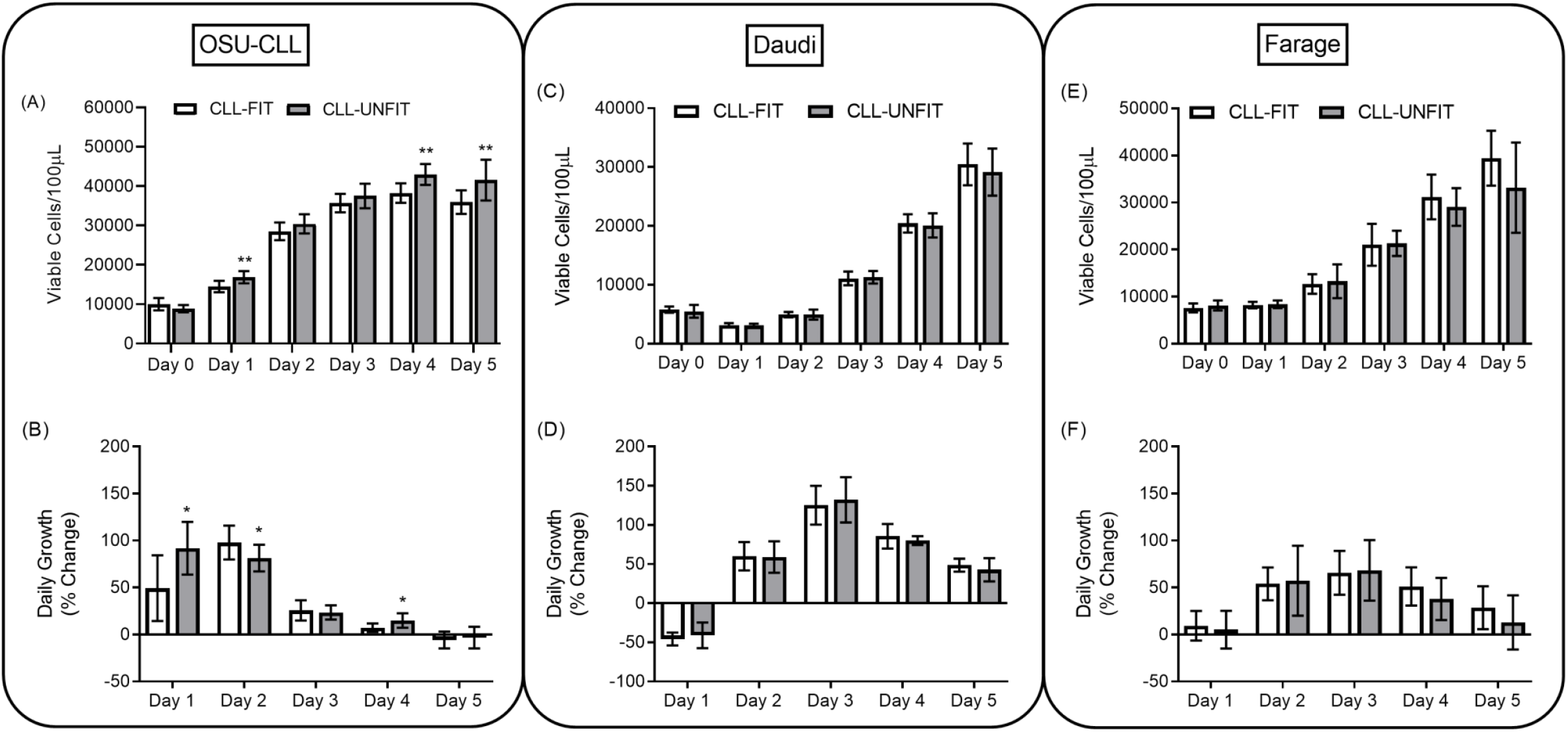
5-Day cell growth in the presence of complete media + 10% autologous plasma from CLL-FIT and CLL-UNFIT. (A) OSU-CLL viable cell counts each day; (B) OSU-CLL viable percentage change from previous day; (C) Daudi viable cell counts each day; (D) Daudi viable percentage change from previous day; (E) Farage viable cell counts each day; (F) Farage viable percentage change from previous day. *p<0.05, **p<0.01 different than CLL-FIT at that corresponding time point. Data are mean ± SD.

**Figure 3.**
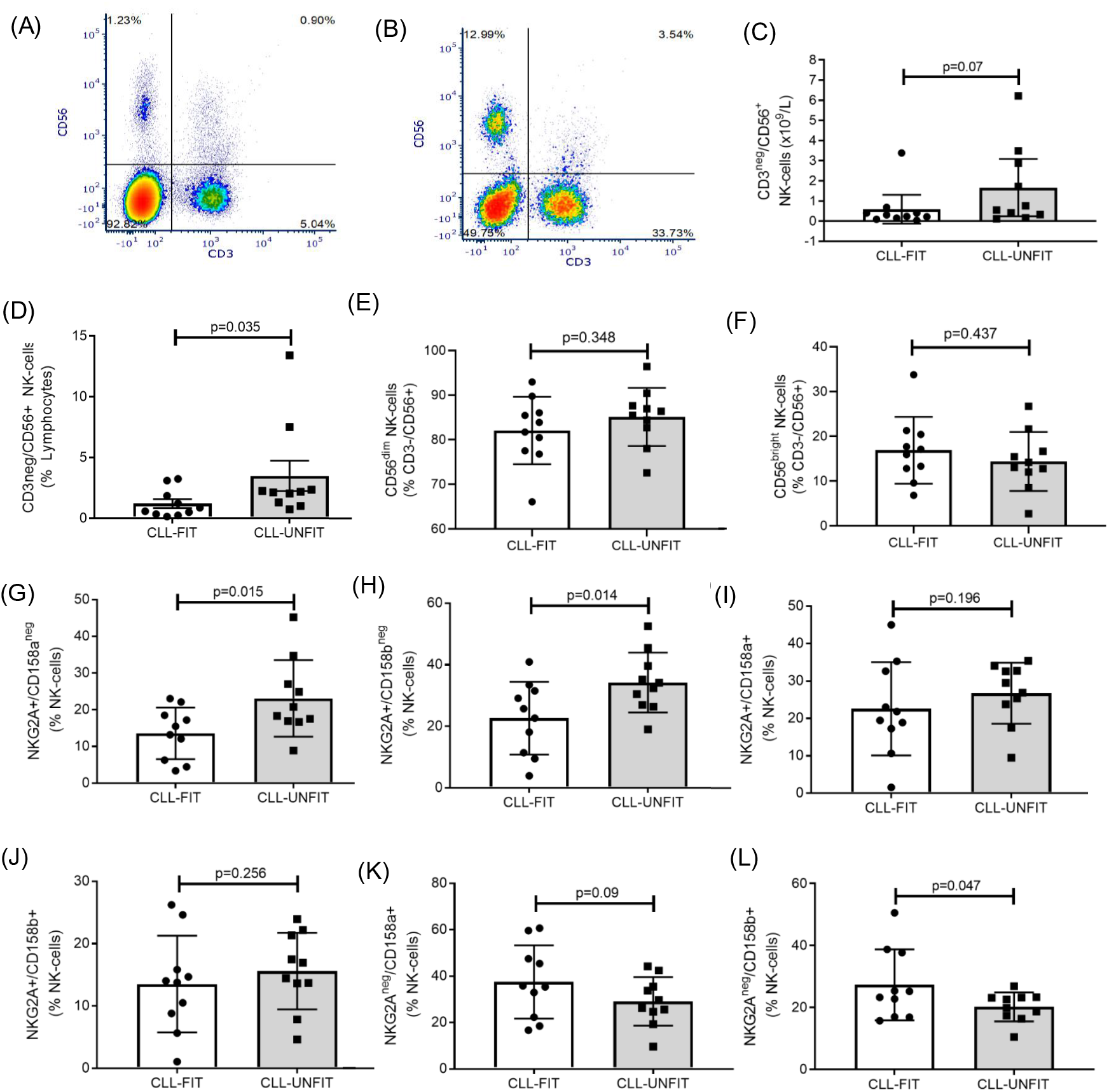
NK-cell similarities and differences between CLL-FIT and CLL-UNFIT. Representative flow cytometry plots of CD3 v CD56 lymphocytes for (A) CLL-FIT and (B) CLL-UNFIT. (C) Absolute numbers of CD3^neg^/CD56+ NK-cells. (D) Percentage of lymphocytes that are CD3^neg^/CD56+ NK-cells. (E) Frequency of CD56^dim^ NK-cells within the NK-cell population. (F) Frequency of CD56^bright^ NK-cells within the NK-cell population. (G) Frequency of least differentiated NKG2A+/CD158a^neg^ NK-cells. (H) Frequency of least differentiated NKG2A+/CD158b^neg^ NK-cells. (I) Frequency of medium differentiated NKG2A+/CD158a+ NK-cells. (J) Frequency of medium differentiated NKG2A+/CD158b+ NK-cells. (K) Frequency of most differentiated NKG2A^neg^/CD158a+ NK-cells. (L) Frequency of most differentiated NKG2A^neg^/CD158b+ NK-cells. Data are mean and 95% CI.

### Exosomal microRNA Profiles

RNAseq revealed a total of 455 distinct exosomal miRNA profiles isolated from patient plasma samples (Supplementary Table 1). To assess similarities and differences in exosomal miRNA profiles between CLL-FIT and CLL-UNFIT patients, we employed a hierarchical clustering method. As shown in Fig. 2A, exosomal miRNA clusters in CLL-FIT vs. CLL-UNFIT differed dramatically, characterized by 32 exosomal miRNAs having a Wald Test *p*-value of ≤0.05 (Supplementary Table 2). Of these, 14 miRNA profiles had ≤-1 or ≥1 log2 fold difference between groups (Fig. 2B). Relative to CLL-UNFIT, CLL-FIT had five miRNAs (miR-378a-3p, miR-32-5p, miR-29c-3p, miR-183-5p, and miR-576-5p) with lower expression (blue dots), and nine miRNAs (miR-130b-5p, miR-1301-3p, miR-4433b-3p, miR-383-5p, miR-328-3p, miR-4433b-5p, miR-324-3p, miR-6772-3p, and miR-1296-5p) with higher expression (red dots). Using miRNA target gene prediction software (miRDB: http://mirdb.org/index.html), we determined which target genes relevant to CLL would be affected by the differential expression of exosomal miRNAs (Supplementary Table 3). There was a pattern for the miRNAs expressed more in CLL-UNFIT to target NOTCH, BCL2, and cyclin signaling.

**Figure 2.**
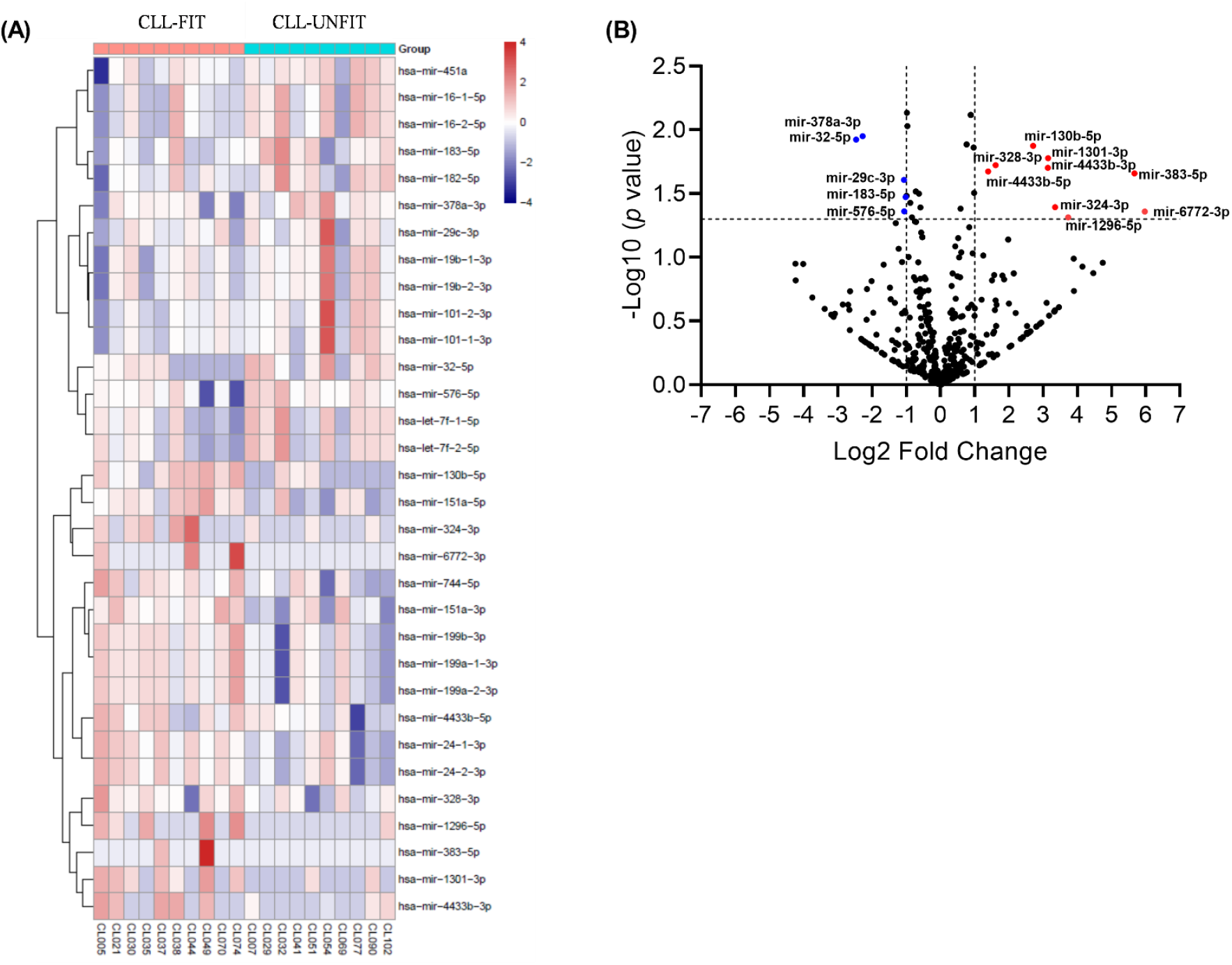
Differential expression of plasma exosomal miRNAs between CLL-FIT and CLL-UNFIT. (A) Heat map with dendrogram illustrating correlation distance with complete linkage hierarchal clustering for the 32-exosomal miRNAs differentially expressed when comparing CLL-FIT to CLL-UNFIT. Values represent the z-score normalized miRNA counts per million reads. Colors represent the level of miRNA expression; with dark red being, the highest expression and dark blue the lowest expression. (B) Volcano plot of differentially expressed miRNAs in the CLL-FIT vs. CLL-UNFIT. The *p*-value in −log10 scale is plotted against the log2 fold change for each miRNA, with each circle denoting an individual miRNA. The miRNAs with Wald Test p-values ≤0.05 and log2 fold change ≥1 or ≤-1 are represented by red circles (higher expression in CLL-FIT) or blue circles (higher expression in CLL-UNFIT). Nine exosomal miRNAs were higher and five were lower in the CLL-FIT compared to CLL-UNFIT group.

### NK-Cell Immunophenotype

Fig. 3A (CLL-FIT) and Fig. 3B (CLL-UNFIT) show representative flow cytometry plots for the frequencies of CD3^neg^/CD56+ NK-cells on total lymphocytes. CLL-FIT patients had a trend for lower absolute numbers of NK-cells (Fig. 3C: *p=*0.07) and lower frequencies of CD3^neg^/CD56+ NK-cells (Fig. 3D; *p=*0.035). Groups were similar for the frequencies of CD56^dim^ (Fig. 3E: *p=*0.348) and CD56^bright^ (Fig. 3F: *p=*0.437) NK-cells. CLL-FIT had a lower frequency of Low-Differentiated NKG2A+/CD158a^neg^ (Fig. 3G: *p=*0.015) and NKGA2A+/CD158b^neg^ (Fig. 3H: *p=*0.014) NK-cells. We noted no differences for Medium-Differentiated NKG2A+/CD158a+ (Fig. 3I: *p=*0.196) or NKG2A+/CD158b+ (Fig. 3J: *p=*0.256) NK-cells. CLL-FIT had a higher frequency of Terminally-Differentiated NKG2A^neg^/CD158b+ (Fig. 3L: *p=*0.047) but not NKG2A^neg^/CD158a+ (Fig. 3K: *p=*0.09) NK-cells. Of the NK-cell surface marker expression (MFI), only CD158a was higher in CLL-FIT (257 ± 32 v 229 ± 24, *p*=0.046), with no differences for CD158b (257 ± 113 v 217 ± 39, *p*=0.301), NKG2A (769 ± 425 v 762 ± 134, *p*=0.960), NKG2C (299 ± 58 v 264 ± 22, *p*=0.093), NKG2D (1293 ± 220 v 1331 ± 133, *p*=0.651), or CD244 (141 ± 48 v 120 ± 5, *p*=0.190), data not shown.

### NMR Measured Lipids, Lipoproteins (LipoProfile^®^), and Inflammatory Profiles

Table 2 shows group differences for pertinent NMR measures. CLL-FIT had lower GlycA concentrations (*p=*0.014) and a lower lipoprotein derived insulin resistance score (LP-IR: *p=*0.007). CLL-FIT had lower concentrations of total triglycerides (*p=*0.025) and triglyceride rich lipoproteins (TRLs; *p=*0.016). CLL-FIT had lower total levels of TRL particles (TRLP: *p=*0.046), large TRLP (*p=*0.011) and very small TRLP (*p=*0.044). CLL-FIT had smaller mean TRL particle size (*p=*0.027). CLL-FIT had lower levels of TRL cholesterol (*p=*0.034), higher HDL cholesterol (*p=*0.040), and higher concentrations of large HDL particles (*p=*0.043). Complete NMR analyses are presented in Supplementary Table 4.

**Table 2:** Select plasma NMR measurements of systemic inflammation, insulin resistance, lipids, and lipoproteins.

|  | CLL-FIT |  | CLL-UNFIT |  | p-Value | 95% C.I. | Effect Size (d) |
| --- | --- | --- | --- | --- | --- | --- | --- |
| **<u>Inflammation</u>** |  |  |  |  |  |  |  |
| GlycA (μmol/L) | 354.7 | ± 31.1 | 442.0 | ± 97.0 | **0.014** | -155.0, -19.6 | 1.21 |
| **<u>Insulin Resistance</u>** |  |  |  |  |  |  |  |
| LP-IR (1-100) | 33.7 | 18.1 | 65.7 | 22.2 | **0.007** | -54.3, -9.7 | 1.58 |
| **<u>Triglycerides</u>** |  |  |  |  |  |  |  |
| Total Triglycerides (mg/dL) | 101.4 | ± 40.6 | 165.2 | ± 72.0 | **0.025** | -118.7, 8.9 | 1.09 |
| TRL Triglycerides (mg/dL) | 78.1 | ± 38.8 | 143.4 | ± 67.7 | **0.016** | -117.1, -13.5 | 1.18 |
| Total TRLP (nmol/L) | 127.6 | ± 58.4 | 189.8 | ± 70.8 | **0.046** | -123.2, -1.2 | 0.96 |
| Large (nmol/L) | 2.5 | ± 3.4 | 8.9 | ± 6.2 | **0.011** | -11.1, -1.7 | 1.28 |
| Very Small (nmol/L) | 73.4 | ± 38.4 | 124.4 | ± 63.7 | **0.044** | -100.4, -1.6 | 0.97 |
| **<u>Cholesterol (mg/dL)</u>** |  |  |  |  |  |  |  |
| Total Cholesterol | 194.4 | ± 29.1 | 195.4 | ± 39.4 | 0.949 | -33.5, 31.5 | 0.03 |
| TRL Cholesterol | 22.5 | ± 10.8 | 34.9 | ± 13.3 | **0.034** | -23.8, -1.0 | 1.02 |
| LDL Cholesterol | 106.3 | ± 25.7 | 105.5 | ± 29.7 | 0.949 | -25.3, 26.9 | 0.03 |
| HDL Cholesterol | 65.5 | ± 10.0 | 54.8 | ± 11.6 | **0.040** | 0.5, 20.9 | 0.99 |
| **<u>HDL-P (μmol/L)</u>** |  |  |  |  |  |  |  |
| Total | 24.7 | ± 2.4 | 23.8 | ± 4.2 | 0.559 | -2.3, 4.1 | 0.26 |
| Large | 3.4 | ± 1.4 | 1.9 | ± 1.6 | **0.043** | 0.1, 2.9 | 1.00 |
| **<u>Lipoprotein Size (nm)</u>** |  |  |  |  |  |  |  |
| TRL | 43.7 | ± 4.7 | 50.3 | ± 7.2 | **0.027** | -12.3, -0.9 | 1.09 |
| LDL | 20.8 | ± 0.4 | 20.4 | ± 0.4 | 0.059 | 0.02, 0.8 | 1.00 |
| HDL | 9.2 | ± 0.4 | 8.8 | ± 0.5 | 0.073 | -0.03, 0.8 | 0.88 |

### Correlations Between miRNAs, Immune Cells and Lipids

CLL-UNFIT patients had higher expression of miR-29c that was associated with higher WBC (r=.558, *p*=0.011) and lymphocyte counts (r=.555, *p*=0.011). CLL-FIT patients had higher expression of miR130b-5p that was associated with lower absolute counts of CD56^bright^ (r=-.493, *p*=0.027) and a trend for CD56^dim^ (r=-.434, *p*=0.056) NK-cells. Additionally, higher expression of miR-130b was associated with lower frequencies of low-differentiated NKG2A+/CD158a^neg^ (r=-.544, *p*=0.013) and highly differentiated mature NKG2A+/CD158b^neg^ NK-cells (r=-.461, *p*=0.041). Higher expression of miR-130b was associated with higher levels of large HDL particles (r=.489, *p*=.029), lower insulin resistance (r=-.498, *p*=0.025), lower large TRLP levels (r=-.497, p-0.026) and smaller TRL lipoprotein size (r=-.451, *p*=0.046). Finally, higher expression of miR-4433b-3p was associated with higher frequencies of highly differentiated NKG2A^neg^/CD158a+ (r=.459, p=0.042) and a trend for higher NKG2A^neg^/CD158b+ NK-cells (r=.390, p=0.089).

## Discussion

For the first time, to our knowledge, we examined the role of physical fitness on *in vitro* malignant cell line growth in older adults with treatment naïve chronic lymphocytic leukemia. Incubation of patient derived plasma with cell lines was associated with differential growth patterns of a CLL-like cell line (OSU-CLL), but not related lymphoma cell lines (Daudi and Farage) suggesting that components in the plasma are specific for the growth of CLL-like cells. Using next generation sequencing, we identified 32 distinct exosomal miRNA profiles with significantly different expression profiles, of which 14 were differentially expressed with a log2 fold difference ≥1 or ≤-1. Many of the exosomal miRNAs that were higher in CLL-UNFIT patients’ plasma were associated with promoting tumor and immune suppression suggesting CLL cells could be removing them from cells in exosomes. Higher fitness was associated with a greater frequency of mature NKG2A^neg^/KIR+ NK-cells. We also identified miRNAs that are associated with lipid homeostasis – a major source for energy and cell signaling unique to CLL cells [44, 45]. Using NMR spectroscopy, we discovered that higher fitness is associated with higher HDL cholesterol, lower triglyceride species (including large and very small triglyceride rich lipoprotein particle levels), triglyceride rich lipoprotein cholesterol levels, and smaller triglyceride rich lipoprotein particle sizes. This cross-sectional analysis for the first time, to our knowledge, shows that physical fitness and function may play a role in the underlying biology of CLL.

### Cell Growth in Autologous Plasma

Our study is the first that we are aware of to demonstrate that serological differences in physically fit CLL patients are associated with slower *in vitro* growth of the CLL-like cell line OSU-CLL. Higher physical fitness is associated with optimal nutritional intake and greater exposure to physical activity and exercise. Following a single bout of exercise in healthy humans, sera/plasma reduces the growth of colorectal, breast, and prostate cancer cell lines, suggesting that exercise increases circulating factors capable of slowing the growth of tumor cells [19, 20, 46]. In mice, 30-days of exercise training reduced lung, melanoma, and liver tumor burden through an IL-6 sensitive and NK-cell mediated mechanism [47]. After each bout of exercise, there are transient increases then decreases upon exercise completion of circulating concentrations of IL-6, and other pro-inflammatory, immunoregulatory cytokines [19, 20, 46-48]. However, resting (i.e. no acute exercise) sera from breast cancer survivors did not alter the growth of breast cancer cell lines following a 6-month exercise intervention [19]. Exercise training reduced circulating concentrations of IL-6 and TNFα, suggesting a more complex role of inflammatory mediators that affects the growth of cell lines *in vitro* [19]. Contrary to findings by Dethlefsen *et al*. [19], we observed lower levels of the chronic inflammatory marker GlycA in CLL-FIT patients and slower *in vitro* cell growth. GlycA is robustly modifiable by exercise training and is lower in those with higher cardiorespiratory fitness [49, 50]. As such, the role inflammation plays on cancer cell growth is complex, and the inflammatory response to exercise which promotes higher physical fitness are equally complex in the cancer setting.

### Potential Circulating Factors Associated With Tumor Cell Growth In Vitro

In an attempt to understand reduced OSU-CLL growth by CLL-FIT plasma, we assessed mediators with known roles in CLL pathophysiology that might be modified by increasing physical fitness. Among the many potential mediators, two include lipids/lipoproteins [44, 45] and exosomal miRNAs [51, 52].

#### Exosomal miRNAs

The role of both intracellular and extracellular (i.e. cell free and packaged in extracellular vesicles) miRNAs in CLL have revealed at least 10-20 miRNAs associated with varying disease characteristics [51-57]. Exosomes are the smallest known extracellular vesicles (50-100 nm) with distinct biochemical properties that carry cellular components, such as proteins, peptides, lipids, mRNA, and miRNAs [58]. miRNAs are small (∼18-28 nt) non-coding RNAs that bind to the specific 3’ UTR of their target mRNA, inducing translational repression, mRNA decapping and deadenylation [59, 60]. miRNAs are transferred from cell to cell in abundance by tumor-derived exosomes and considered as potential disease biomarkers [61, 62]. Exosomes are constitutively secreted in response to B-cell receptor activation and stress by CLL cells. The roles of these miRNAs include promotion of CLL cell survival by (a) altering transcription in the CLL cell, (b) altering functions of cells of the tumor microenvironment, and (c) altering effector functions of immune cells such as CD8+ T-cells and NK-cells, and thus promoting immune evasion and chronic immune suppression [61, 63, 64].

#### miRNAs and Physical Fitness

Thus far, only one study that we are aware of has assessed differences in exosomal miRNA expression between sedentary (N=5) and physically fitter (N=5) older adults [65]. In the fitter group, Nair and colleagues found that exosomal miR-486-5p, miR-215-5p, and miR-941were upregulated, but that miR-151b was downregulated compared to that in the control group – none of which we observed significant differences for in our study [65]. Although the ages of the Nair study participants were similar to those in our study, their subjects were all male and differences between their groups and our groups for fitness were less (Nair = 37% v our 27%) and for group’s BMI was greater (Nair = 10% v our 26%). With this, and the addition of CLL, it is not possible to compare our results other than to appreciate that physical fitness is likely associated with differential expression of exosomal miRNAs in older adults. As such, we discuss our results in terms of known tumor promotion and immune suppression strategies of differentially expressed miRNAs.

#### miRNAs and Cell Survival

Compared to healthy adults, CLL patient exosomes have higher expression of miR-29c that is associated with elevated white blood cell and lymphocyte counts [52, 63, 66]. Here, we show that CLL-UNFIT patients have higher expression of exosomal miR-29c, and that higher expression is associated with higher WBC and lymphocyte counts. In patients with unfavorable prognostic markers, miR-29c is downregulated inside (rather than outside in exosomes) CLL cells [67]. Compared to healthy donor B-cells, normal B-cells from CLL patients have similar expression of miR-29 [63]. This suggests that CLL cells are preferentially expelling miR-29 inside exosomes. miR-29c targets multiple NOTCH1, BCL2 and cyclin related mRNAs, post-transcriptionally repressing translation and impacting regulatory control of cell apoptosis, growth and proliferation. Additionally, miR-15a (p=0.0529) and miR-16-1 (p=0.0304) were both approximately 2-fold lower in CLL-FIT patient exosomes. These exosomes are two of the first to be identified as regulators of CLL cell survival [68]. Both promote BCL2 repression and induce apoptosis in leukemia cell lines, and as such are downregulated inside CLL cells. CLL cells are likely expelling miR-29c, and other negative regulators, to promote their survival and growth. Although lower physical fitness might be associated with more secretion of miR-29c from CLL cells, it is plausible that higher fitness is causing more *in vivo* uptake of exosomes containing miR-29c. However, it is not clear from which cells these miRNAs would originate.

#### miRNAs and Immune Function

Many tumor-originating exosomal miRNAs function by suppressing the normal immune system’s ability to recognize and kill tumor cells [69, 70]. Of these, exosomal miR-29c, miR-378, miR-183 and miR-130b were differentially expressed in our two groups. miR-29c, miR-378, and miR-183 each target NK-cell and some CD8+ T-cell functions, including IFNγ production, granzyme B formation and activatory receptor expression [71-75]. As such, it is plausible that more CLL derived exosomal miR-29c, miR-378, and miR-183 are being delivered to more NK-cells and CD8+ T-cells observed in CLL-UNFIT patients. Higher miR-130b, the only one with higher expression in CLL-FIT patients, correlated with lower absolute numbers and frequencies of NK-cells – a characteristic noted in healthy adults compared to CLL [76, 77]. Although it is unclear if miR-130b influences NK-cells, it has been shown to disrupt the CLL mediated polarization of immuno-suppressive macrophages [78]. In mice, miR-130b promotes macrophage differentiation into a more favorable M1 phenotype, while sera from CLL patients has been noted to result in induction of monocyte differentiation towards an M2 macrophage phenotype [78, 79]. Further studies are required to elucidate whether higher physical fitness is promoting immune surveillance by selectively increasing (i.e. miR-130b) and lowering (i.e. miR-29c, miR-378 and miR-183) exosomal miRNAs that influence effector immune cell functions.

Of the remaining miRNAs expressed more in CLL-FIT patient exosomes, less is known for CLL. In chronic myelogenous leukemia (CML) cells, increasing expression of miR-328-3p and miR-4433b reduces blast cell survival and impairs cell growth [80, 81]. Exosomal miR-32-5p activates the PI3K/Akt pathway and suppresses PTEN in solid tumor cells [82].

Taken together, differential expression of exosomal miRNAs plays an important role in the pathophysiology of CLL [51, 56]. We provide a novel insight into possible mechanisms by which physical fitness might influence CLL biology. Most of the exosomal miRNAs that are higher in CLL-UNFIT target NOTCH, Cyclins, and BCL2 signaling (Supplementary Table 3). These pathways are typically overexpressed in CLL and offer favorable prognosis when suppressed or inhibited. As such, CLL-UNFIT cells appear to be selectively expelling tumor-suppressing miRNAs in exosomes [83], and these exosomes likely do not bind efficiently with malignant cells or deliver their contents to neighboring tumor cells [61]. This is particularly evident given that CLL-UNFIT plasma is associated with increased growth of a CLL-like cell line while having similar mean CLL absolute cell numbers as CLL-FIT. Consequently, understanding the role of physical fitness on exosomal miRNA functions may lead to novel understandings of CLL progression and immune evasion.

#### Lipid/Lipoprotein Homeostasis

There are much data to suggest that in overweight and obese older adults, regular exercise and/or weight loss can affect health outcomes by improving profiles of lipids and lipoproteins [84]. Here, CLL-FIT patients had higher levels of HDL cholesterol, and lower triglycerides and triglyceride rich lipoproteins – levels consistent with better physical fitness [85]. However, compared to healthy adults, CLL levels of cholesterol, HDL and LDL are lower [86, 87]. In addition, these lipids are lowered more as disease stage increases [86]. McCaw and colleagues, using an *in vitro* pseudofollicle model of CLL proliferation, suggest that lipids and LDLs are preferentially utilized by CLL cells for cell signaling functions [44]. Specifically, LDL mediated amplification of STAT3 phosphorylation and cholesterol dense lipid rafts uniquely improve CLL cell survival and increase proliferation [44]. As such, preventing this CLL-lipid process should negatively regulate lipid metabolism in CLL cells. Indeed, lipid-lowering medications confer longer time to first treatment and improved CLL patient survival [45, 88]. Interestingly, we show here that higher exosomal miR-130b expression is associated with better lipid profiles suggesting that miR-130b has a role in CLL lipid homeostasis. Indeed, miR-130b is a potential regulator of lipid homeostasis, targeting lipid oxidation and LDL receptor expression [89, 90]. That said, dyslipidemia and hypercholesterolemia also likely have complex roles in the pathogenesis of CLL, but these are not fully understood [91, 92]. Lowering these lipids and lipoproteins by either medications, diet, or exercise (before CLL cells can utilize them) could modulate CLL cell growth, and warrants further investigation.

#### The Immune System

Higher physical fitness and regular physical activity in older age maintains and improves important immune functions associated with reduced risks of infections, cancer, and chronic diseases [93-97]. Indeed, older fitter and healthier adults have T-cell and B-cell characteristics more similar to adults many years younger [98, 99], while NK-cells have improved function in those with increased physical fitness [24, 47]. Compared to healthy adults, newly diagnosed and treatment naive CLL is associated with higher absolute numbers of CD4+ and CD8+ T-cell, and NK-cells [76, 77]. In CLL, NK-cell and T-cell expansion is driven by repeated cycles of replication due to increased infections [77]. CLL NK- and T-cells are characterized by increased inhibitory receptor expression (e.g. PD-1, Tim3, and CTLA-4) indicative of an “exhausted” phenotype, contributing to poor resolution of infections [100]. We show that CLL-FIT patients have lower frequencies of NK-cells, and lower absolute numbers of CD8+ and CD4+ T-cells (albeit non-significantly), and NK-cells—all similar to observations in healthy adults [76, 77]. Further, CLL reduces NK-cell tumor cytotoxicity, reduces expression of activatory NK-cell receptors, and increases inhibitory NK-cell receptor expression [76, 101]. We show in the current study that CLL-FIT patients have a lower frequency of inhibitory positive/activatory negative (i.e. NKG2A^+^/KIR^neg^) NK-cells and higher frequency of fully competent mature inhibitory negative/activatory positive (i.e. NKG2A^neg^/KIR^+^) NK-cells that are better capable of recognizing and killing tumor cells [102]. Since we did not observe absolute number differences in malignant cells, it may be that CLL NK-cells are less responsive to physical fitness. Alternatively, it may be that over several years, CLL cells from CLL-FIT patients have not been exposed to as many infections that drive increased immune cell numbers. Another possible explanation for this could be that CLL-UNFIT patients have more CLL cells residing in primary and secondary lymphoid tissues. Although our groups were similarly matched for disease stage, increasing lymphoid tissue size is a characteristic of disease progression due to the accumulation of malignant CLL cells. Unfortunately, we were unable to determine whether there were lymph node or splenic size differences in our groups, but given the variation in absolute lymphocyte counts in the blood it is likely that there is also large variation in the lymphoid tissue. A potential way to assess this is through measuring lymphocyte counts before and immediately after an acute bout of moderate-vigorous intensity exercise. An acute bout of exercise temporarily increases circulating blood lymphocyte counts, before counts return to normal in the hours following exercise completion [93]. Lymphocytes move into the blood from the marginated pool and secondary lymphoid tissues allowing quantification of total body lymphocyte counts, rather than just those in the blood [103]. Perry *et al*. showed that in four CLL patients’ lymphocyte counts increased by 40% following exercise (resting: 18.7 ± 6.6 ×10^6^/mL to immediately post exercise: 26.3 ± 11.4 ×10^6^/mL)[104]. With no changes for CD4+ T-cell counts, the majority of the increase was CD8+ T-cells, NK-cells and CD19+ B-cells. Further studies are required to determine the role of physical fitness on normal immune cell phenotype and function in CLL.

## Limitations and Future Directions

Our study is not without limitations. Our sample size was small and as such limits our interpretations of the data. Larger sample sizes should be assessed to confirm our findings. We used autologous plasma and fetal bovine serum (control) for tumor cell incubation assays. We did this to ensure the composition of the plasma used for NMR and RNAseq were similar to the cell assays. Although we tried to encourage all patients to perform their best and to exert maximal efforts in the physical assessments, we can never be certain that this was done, but differences in other functional measures did not reflect this. With significant differences in SPPB and better (albeit non-significant) values for TUG and normalized grip we are confident our groups are representative of the fitter and least fit CLL patients. We did not explore comprehensive phenotypes or functions of T-cells or B-cells. As such, it remains unclear whether physical fitness is associated with distinct subsets of T- and B-cells that may be critical to CLL progression. Whether the differences in NK-cells reflect better tumor control mechanisms is also unclear. There are two important future directions for our research: (a) confirm our findings in a larger cohort with more comprehensive physiological and biological phenotyping, with inclusion of more functional measures of tumor control, including culture of autologous malignant B-cells, and (b) attempt to change the physical fitness of CLL patients and determine which of our biological measures changes accordingly.

## Conclusions

In treatment naïve CLL, the CLL-like cell line (OSU-CLL) cultured *in vitro* with plasma from patients with higher physical fitness results in less growth/survival as compared to cells cultured in plasma from CLL patients with lower physical fitness. Higher fitness is associated with greater frequency of mature NK-cells, lower triglyceride concentrations, and differentially expressed miRNAs in plasma exosomes. The combination of these factors likely reflects improved health outcomes associated with being physically fit. Importantly, we noted that physical fitness is not associated with worsening CLL outcomes, and suggest that being fitter imparts a reduced risk of diabetes (lower insulin resistance), less cardiovascular disease (lower GlycA inflammation), and potentially less secondary malignancies and infections (higher frequency of mature NK-cells).

## Data Availability

The datasets generated during the present study are not publicly available, owing to the risk of disclosure or deduction of private individual information, but they are available from the corresponding author on reasonable request.

## Author Contributions

DBB, DMB, JBW and AS conceived and designed the study and experimental approach. EG completed the NMR spectroscopy and data analysis.MAD, DKT, and TS identified and completed physical testing on the patients. DBB and GM performed the cell growth assay. ND and DC completed the RNAseq and analysis. JS, JE, KW helped design and analyze the flow cytometry data. All authors contributed to critical revisions and approval of the final manuscript.

## Acknowledgements

The research team acknowledges the support of the Duke Cancer Center and the support provided by nurses and clinical staff during the execution of the study. We also acknowledge Prof. John Byrd from the Ohio State University for providing us with the OSU-CLL cell line. Finally, we appreciate the participants who kindly gave up their time to take part in this study.

## Abbreviations

6MWT: 6-minute walk test
BCAA: branch chain amino acid
CBC: complete blood count
CD: cluster of differentiation
CLL: Chronic Lymphocytic Leukemia
CLL-IPI: Chronic Lymphocytic Leukemia – International Prognostic Index
DMSO: Dimethyl sulfoxide
DPBS: Dulbecco’s phosphate buffered saline
EBV: Epstein Barr Virus
EDTA: Ethylenediaminetetraacetic acid
eVO2peak: estimated peak volume of oxygen consumption
FBS: fetal bovine serum
FFA: Free Fatty Acids
FMO: flow minus one
HDL: high-density lipoprotein
ICAM-1: Intercellular Adhesion Molecule 1
IFN: interferon
IGHV: immunoglobulin heavy chain variable region genes
IL: interleukin
IPAQ: Incidental and Planned Activity Questionnaire
KIR: Killer immunoglobulin-like receptor
LDL: low-density lipoprotein
LP-IR: Lipoprotein Insulin Resistance Index
miRNA: micro-RNA
NK-cell: Natural Killer Cell
NKG2: CD94/NK group 2 member
NMR: nuclear magnetic resonance
OSU-CLL: Ohio State University-Chronic Lymphocytic Leukemia
PBMC: peripheral blood mononuclear cell
PI: propidium iodide
RNA: ribonucleic acid
RPMI: Roswell Park Memorial Institute
SBAS: Stanford Brief Activity Survey
SPPB: short physical performance battery
STAT: Signal transducer and activator of transcription
TGFβ: Transforming growth factor β
TNFα: tumor necrosis factor α
TRL: triglyceride rich lipoprotein
TRLP: triglyceride rich lipoprotein particle
TUG: timed-up and go
UMI: Unique Molecular Indices

---

**Supplementary Table 1.** Complete List of miRNAs identified.

| miRNA | log2 Fold Change<br>(CLL-FIT/CLL-UNFIT) | Log2<br>Fold<br>(S.E.) | Wald Test<br>Statistic | Wald Test<br>p value |
| --- | --- | --- | --- | --- |
| hsa-mir-101-2-3p | -0.983875786 | 0.36716 | -2.679722209 | 0.0074 |
| hsa-mir-199a-1-3p | 0.88129529 | 0.33034 | 2.667857739 | 0.0076 |
| hsa-mir-199a-2-3p | 0.88129529 | 0.33034 | 2.667857739 | 0.0076 |
| hsa-mir-199b-3p | 0.88129529 | 0.33034 | 2.667857739 | 0.0076 |
| hsa-mir-101-1-3p | -0.971037046 | 0.37353 | -2.599613693 | 0.0093 |
| hsa-mir-378a-3p | -2.282514775 | 0.90012 | -2.535796826 | 0.0112 |
| hsa-mir-32-5p | -2.470054298 | 0.98256 | -2.513902987 | 0.0119 |
| hsa-mir-24-1-3p | 0.758239422 | 0.30543 | 2.482527222 | 0.013 |
| hsa-mir-24-2-3p | 0.758239422 | 0.30543 | 2.482527222 | 0.013 |
| hsa-mir-130b-5p | 2.702963742 | 1.09236 | 2.474433631 | 0.0133 |
| hsa-mir-744-5p | 0.966822005 | 0.39263 | 2.462433206 | 0.0138 |
| hsa-mir-1301-3p | 3.145610151 | 1.31455 | 2.392918164 | 0.0167 |
| hsa-mir-328-3p | 1.608130759 | 0.68509 | 2.347339234 | 0.0189 |
| hsa-mir-4433b-3p | 3.13815109 | 1.3475 | 2.328873325 | 0.0199 |
| hsa-mir-4433b-5p | 1.388521901 | 0.60249 | 2.304643691 | 0.0212 |
| hsa-mir-383-5p | 5.668882623 | 2.47465 | 2.29078449 | 0.022 |
| hsa-mir-29c-3p | -1.069640841 | 0.47632 | -2.245652119 | 0.0247 |
| hsa-mir-16-1-5p | -0.727284521 | 0.33597 | -2.164725857 | 0.0304 |
| hsa-mir-16-2-5p | -0.727284521 | 0.33597 | -2.164725857 | 0.0304 |
| hsa-mir-151a-5p | 0.97675396 | 0.45355 | 2.153560989 | 0.0313 |
| hsa-let-7f-1-5p | -0.635424076 | 0.29586 | -2.147697168 | 0.0317 |
| hsa-mir-19b-1-3p | -0.998943501 | 0.46834 | -2.132966674 | 0.0329 |
| hsa-mir-19b-2-3p | -0.994104722 | 0.46627 | -2.132056046 | 0.033 |
| hsa-mir-183-5p | -1.020194315 | 0.48043 | -2.123496223 | 0.0337 |
| hsa-mir-451a | -0.885223579 | 0.42546 | -2.080613285 | 0.0375 |
| hsa-mir-324-3p | 3.34659127 | 1.63312 | 2.049206459 | 0.0404 |
| hsa-let-7f-2-5p | -0.590170504 | 0.2884 | -2.046353137 | 0.0407 |
| hsa-mir-151a-3p | 0.582577816 | 0.28579 | 2.038501319 | 0.0415 |
| hsa-mir-576-5p | -1.062177535 | 0.52628 | -2.018270367 | 0.0436 |
| hsa-mir-6772-3p | 5.973170484 | 2.96149 | 2.016948525 | 0.0437 |
| hsa-mir-182-5p | -0.837563539 | 0.4247 | -1.97212436 | 0.0486 |
| hsa-mir-1296-5p | 3.726897256 | 1.89014 | 1.971759554 | 0.0486 |
| hsa-mir-142-5p | -0.752124146 | 0.38777 | -1.939613632 | 0.0524 |
| hsa-mir-15a-5p | -0.71711871 | 0.37039 | -1.936142158 | 0.0529 |
| hsa-mir-96-5p | -1.314855247 | 0.68199 | -1.927956332 | 0.0539 |
| hsa-mir-1307-3p | 0.837645679 | 0.44225 | 1.894057784 | 0.0582 |
| hsa-let-7g-5p | -0.569977944 | 0.30771 | -1.852294927 | 0.064 |
| hsa-let-7a-1-5p | -0.531267809 | 0.29274 | -1.814835503 | 0.0695 |
| hsa-let-7a-2-5p | -0.531267809 | 0.29274 | -1.814835503 | 0.0695 |
| hsa-let-7a-3-5p | -0.531267809 | 0.29274 | -1.814835503 | 0.0695 |
| hsa-mir-339-5p | 0.51066174 | 0.28264 | 1.806736469 | 0.0708 |
| hsa-mir-199a-1-5p | 1.975049998 | 1.09976 | 1.795891286 | 0.0725 |
| hsa-mir-199a-2-5p | 1.975049998 | 1.09976 | 1.795891286 | 0.0725 |
| hsa-mir-191-5p | 0.426561535 | 0.24535 | 1.738602443 | 0.0821 |
| hsa-mir-29a-3p | -1.226522299 | 0.71389 | -1.71808784 | 0.0858 |
| hsa-mir-584-5p | 0.606666997 | 0.35924 | 1.688734072 | 0.0913 |
| hsa-mir-139-3p | 0.936260741 | 0.55753 | 1.679300781 | 0.0931 |
| hsa-mir-140-5p | 1.24689876 | 0.75116 | 1.659954468 | 0.0969 |
| hsa-mir-20b-5p | -0.934883701 | 0.56733 | -1.647852597 | 0.0994 |
| hsa-mir-223-3p | 0.541384276 | 0.32938 | 1.643647946 | 0.1002 |
| hsa-mir-3064-5p | 3.89556136 | 2.38492 | 1.633417269 | 0.1024 |
| hsa-mir-500a-3p | -1.126655578 | 0.7024 | -1.604006715 | 0.1087 |
| hsa-mir-10a-5p | -0.651213235 | 0.40674 | -1.601036544 | 0.1094 |
| hsa-mir-331-5p | 4.73880117 | 2.96803 | 1.5966131 | 0.1104 |
| hsa-mir-629-3p | -4.244348122 | 2.67072 | -1.58921359 | 0.112 |
| hsa-mir-664b-3p | -4.017289695 | 2.53324 | -1.585833248 | 0.1128 |
| hsa-mir-1291 | -1.661940698 | 1.0522 | -1.579492975 | 0.1142 |
| hsa-mir-2355-3p | 4.149533168 | 2.65535 | 1.562703966 | 0.1181 |
| hsa-mir-6738-5p | 4.463026457 | 2.97042 | 1.502491583 | 0.133 |
| hsa-mir-133a-1-3p | 2.136473363 | 1.42483 | 1.499454912 | 0.1338 |
| hsa-mir-133a-2-3p | 2.136473363 | 1.42483 | 1.499454912 | 0.1338 |
| hsa-mir-142-3p | 0.349845229 | 0.23339 | 1.498971514 | 0.1339 |
| hsa-mir-483-5p | 1.566411423 | 1.05549 | 1.484057072 | 0.1378 |
| hsa-mir-5187-5p | 1.805291799 | 1.21942 | 1.480456208 | 0.1388 |
| hsa-mir-221-3p | 0.492404648 | 0.33451 | 1.471998366 | 0.141 |
| hsa-mir-143-3p | 0.568444726 | 0.38817 | 1.464415718 | 0.1431 |
| hsa-mir-30e-3p | -0.780048649 | 0.53283 | -1.463969616 | 0.1432 |
| hsa-mir-140-3p | -0.47168392 | 0.32297 | -1.460475962 | 0.1442 |
| hsa-mir-30a-5p | -0.592087769 | 0.40836 | -1.449904436 | 0.1471 |
| hsa-mir-181a-2-5p | -0.440722775 | 0.30479 | -1.445981503 | 0.1482 |
| hsa-mir-664b-5p | 1.857625403 | 1.28671 | 1.443704221 | 0.1488 |
| hsa-mir-660-5p | -0.731350959 | 0.50937 | -1.435785567 | 0.1511 |
| hsa-let-7g-3p | -4.240436069 | 2.95536 | -1.434828286 | 0.1513 |
| hsa-mir-505-5p | 1.504533296 | 1.04949 | 1.433586627 | 0.1517 |
| hsa-mir-1299 | -2.015661936 | 1.41262 | -1.42689401 | 0.1536 |
| hsa-mir-320a | 0.324779735 | 0.23536 | 1.379916189 | 0.1676 |
| hsa-mir-4732-5p | -1.479082901 | 1.08592 | -1.362057497 | 0.1732 |
| hsa-mir-497-5p | -2.155441964 | 1.59736 | -1.349377501 | 0.1772 |
| hsa-mir-484 | -0.603713321 | 0.44876 | -1.345283012 | 0.1785 |
| hsa-mir-93-5p | -0.35842289 | 0.26817 | -1.336566345 | 0.1814 |
| hsa-mir-3164 | 3.898097408 | 2.93199 | 1.329504889 | 0.1837 |
| hsa-let-7b-5p | -0.487144465 | 0.36684 | -1.327949971 | 0.1842 |
| hsa-mir-651-5p | -2.649315881 | 1.99809 | -1.325924204 | 0.1849 |
| hsa-mir-144-3p | -0.70339845 | 0.53144 | -1.323582713 | 0.1856 |
| hsa-mir-27a-3p | -0.687787587 | 0.52055 | -1.321266032 | 0.1864 |
| hsa-mir-21-5p | -0.577539264 | 0.44548 | -1.296437372 | 0.1948 |
| hsa-mir-144-5p | -0.61077933 | 0.48262 | -1.265560243 | 0.2057 |
| hsa-mir-5010-3p | -3.74838597 | 2.96791 | -1.262971665 | 0.2066 |
| hsa-let-7c-5p | 0.441837838 | 0.35343 | 1.25015088 | 0.2112 |
| hsa-mir-3158-1-3p | -1.459726023 | 1.17272 | -1.244738177 | 0.2132 |
| hsa-mir-3158-2-3p | -1.459726023 | 1.17272 | -1.244738177 | 0.2132 |
| hsa-mir-1307-5p | 1.200211251 | 0.96749 | 1.240547475 | 0.2148 |
| hsa-mir-1287-5p | 1.608184763 | 1.30476 | 1.232548282 | 0.2177 |
| hsa-mir-376c-3p | -1.342712173 | 1.1127 | -1.20671296 | 0.2275 |
| hsa-mir-3200-3p | 3.103176477 | 2.57273 | 1.206181625 | 0.2277 |
| hsa-mir-181d-5p | 1.99394513 | 1.66279 | 1.199156094 | 0.2305 |
| hsa-mir-25-3p | -0.3641706 | 0.30582 | -1.19081383 | 0.2337 |
| hsa-mir-570-3p | -2.857004846 | 2.40372 | -1.188576114 | 0.2346 |
| hsa-mir-15b-3p | 0.903633226 | 0.76148 | 1.186675883 | 0.2354 |
| hsa-mir-362-5p | 1.637914238 | 1.38069 | 1.186304002 | 0.2355 |
| hsa-mir-31-5p | -2.703802932 | 2.27987 | -1.185943668 | 0.2356 |
| hsa-mir-30b-5p | -0.607985789 | 0.52084 | -1.167324952 | 0.2431 |
| hsa-mir-212-3p | 3.463673702 | 2.98396 | 1.16076351 | 0.2457 |
| hsa-mir-486-2-5p | -0.493572567 | 0.42601 | -1.158604844 | 0.2466 |
| hsa-mir-491-5p | 0.81073414 | 0.70616 | 1.148089643 | 0.2509 |
| hsa-let-7d-3p | 1.001751966 | 0.87309 | 1.147369578 | 0.2512 |
| hsa-mir-150-3p | -3.388448154 | 2.9711 | -1.140470677 | 0.2541 |
| hsa-mir-203a-3p | -2.667249535 | 2.3567 | -1.131772687 | 0.2577 |
| hsa-mir-6858-5p | 1.584063222 | 1.40565 | 1.126928019 | 0.2598 |
| hsa-mir-5698 | 3.343729312 | 2.97406 | 1.124296149 | 0.2609 |
| hsa-mir-423-3p | 0.336591286 | 0.2994 | 1.124202189 | 0.2609 |
| hsa-mir-1294 | -1.042776142 | 0.93021 | -1.121016389 | 0.2623 |
| hsa-mir-3064-3p | 3.333949929 | 2.98636 | 1.116393602 | 0.2643 |
| hsa-mir-3180-2 | 3.331709622 | 2.98639 | 1.115630111 | 0.2646 |
| hsa-mir-3180-4 | 3.331709622 | 2.98639 | 1.115630111 | 0.2646 |
| hsa-mir-3180-5 | 3.331709622 | 2.98639 | 1.115630111 | 0.2646 |
| hsa-mir-92a-2-3p | -0.449038872 | 0.40295 | -1.114385426 | 0.2651 |
| hsa-mir-766-5p | 3.323130745 | 2.98653 | 1.112706431 | 0.2658 |
| hsa-let-7d-5p | -0.332395837 | 0.30114 | -1.103797346 | 0.2697 |
| hsa-mir-23a-3p | 0.284144706 | 0.2588 | 1.09793917 | 0.2722 |
| hsa-mir-190b | -1.968724688 | 1.79362 | -1.097623767 | 0.2724 |
| hsa-mir-335-3p | -1.029920973 | 0.93958 | -1.096152013 | 0.273 |
| hsa-mir-99b-3p | 2.194749348 | 2.00433 | 1.095003674 | 0.2735 |
| hsa-mir-486-1-5p | -0.429168897 | 0.39309 | -1.091781631 | 0.2749 |
| hsa-mir-130b-3p | -1.127433105 | 1.03524 | -1.089052114 | 0.2761 |
| hsa-mir-3200-5p | -3.094777686 | 2.84561 | -1.0875607 | 0.2768 |
| hsa-mir-361-3p | 0.419272606 | 0.38642 | 1.085020186 | 0.2779 |
| hsa-mir-7704 | -3.197168331 | 2.97314 | -1.075351171 | 0.2822 |
| hsa-mir-7-1-5p | -0.438229113 | 0.40815 | -1.073699792 | 0.283 |
| hsa-mir-3144-3p | 3.168918061 | 2.97653 | 1.064633578 | 0.287 |
| hsa-mir-374b-5p | 0.990212585 | 0.93098 | 1.06362797 | 0.2875 |
| hsa-mir-224-5p | 0.638471575 | 0.60309 | 1.058665143 | 0.2898 |
| hsa-mir-585-3p | -3.141012384 | 2.97379 | -1.056231652 | 0.2909 |
| hsa-mir-128-2-3p | 0.372999753 | 0.35386 | 1.054078666 | 0.2918 |
| hsa-mir-629-5p | 0.428326695 | 0.40673 | 1.053088705 | 0.2923 |
| hsa-mir-340-5p | 0.589314566 | 0.56054 | 1.051338137 | 0.2931 |
| hsa-mir-3150b-3p | -3.125661517 | 2.97397 | -1.051005013 | 0.2933 |
| hsa-mir-4732-3p | -0.903155739 | 0.86624 | -1.04261025 | 0.2971 |
| hsa-mir-28-3p | 0.370397656 | 0.3561 | 1.040152227 | 0.2983 |
| hsa-mir-128-1-3p | 0.353124571 | 0.34101 | 1.035521935 | 0.3004 |
| hsa-mir-363-3p | -0.575093262 | 0.55593 | -1.034476739 | 0.3009 |
| hsa-mir-15b-5p | -0.30785793 | 0.29798 | -1.033151978 | 0.3015 |
| hsa-mir-181a-1-5p | -0.298280161 | 0.29018 | -1.027924817 | 0.304 |
| hsa-mir-92a-1-3p | -0.410013596 | 0.40209 | -1.019708428 | 0.3079 |
| hsa-mir-28-5p | -2.16503415 | 2.12925 | -1.016806371 | 0.3092 |
| hsa-mir-532-5p | -0.360212089 | 0.36018 | -1.000086417 | 0.3173 |
| hsa-mir-7-2-5p | -0.404392394 | 0.40799 | -0.991174288 | 0.3216 |
| hsa-mir-7-3-5p | -0.404392394 | 0.40799 | -0.991174288 | 0.3216 |
| hsa-mir-433-3p | 2.941873896 | 2.99348 | 0.982759389 | 0.3257 |
| hsa-mir-548ac | 2.901867296 | 2.99432 | 0.969123648 | 0.3325 |
| hsa-mir-196b-5p | -0.359039909 | 0.37849 | -0.94860875 | 0.3428 |
| hsa-mir-19a-3p | -0.557186371 | 0.59016 | -0.9441336 | 0.3451 |
| hsa-mir-1-1-3p | 1.622975393 | 1.72304 | 0.941927949 | 0.3462 |
| hsa-mir-1-2-3p | 1.622975393 | 1.72304 | 0.941927949 | 0.3462 |
| hsa-mir-939-5p | 2.524382984 | 2.68058 | 0.941731891 | 0.3463 |
| hsa-mir-548u | 2.812373096 | 2.99628 | 0.938622394 | 0.3479 |
| hsa-mir-185-3p | 2.797473071 | 2.99661 | 0.933544498 | 0.3505 |
| hsa-mir-424-5p | -1.249246606 | 1.38992 | -0.898790946 | 0.3688 |
| hsa-mir-107 | -0.300912229 | 0.33522 | -0.8976575 | 0.3694 |
| hsa-mir-1288-3p | -2.656902516 | 2.98059 | -0.891402027 | 0.3727 |
| hsa-mir-320b-1 | 0.670591588 | 0.76095 | 0.881260057 | 0.3782 |
| hsa-mir-320b-2 | 0.670591588 | 0.76095 | 0.881260057 | 0.3782 |
| hsa-mir-3150a-5p | 2.627042004 | 2.99993 | 0.875700119 | 0.3812 |
| hsa-mir-30e-5p | -0.294007045 | 0.33702 | -0.872376567 | 0.383 |
| hsa-mir-206 | 1.569659152 | 1.79942 | 0.872315899 | 0.383 |
| hsa-mir-324-5p | 0.573213858 | 0.6609 | 0.867328367 | 0.3858 |
| hsa-mir-509-1-3p | 2.584370053 | 3.00063 | 0.861276983 | 0.3891 |
| hsa-mir-509-2-3p | 2.584370053 | 3.00063 | 0.861276983 | 0.3891 |
| hsa-mir-509-3-3p | 2.584370053 | 3.00063 | 0.861276983 | 0.3891 |
| hsa-mir-26b-5p | -0.203933619 | 0.23689 | -0.860871156 | 0.3893 |
| hsa-mir-4746-5p | 2.523122144 | 2.94426 | 0.856963108 | 0.3915 |
| hsa-mir-122-5p | 0.420232104 | 0.49261 | 0.853066388 | 0.3936 |
| hsa-mir-222-3p | -0.446158492 | 0.5258 | -0.848539811 | 0.3961 |
| hsa-mir-3605-3p | 1.535718265 | 1.81091 | 0.848038263 | 0.3964 |
| hsa-mir-4747-5p | 2.531115247 | 3.00152 | 0.843278364 | 0.3991 |
| hsa-mir-636 | 2.531115247 | 3.00152 | 0.843278364 | 0.3991 |
| hsa-mir-18a-5p | -0.655981756 | 0.78577 | -0.834822631 | 0.4038 |
| hsa-mir-326 | -1.87474568 | 2.24649 | -0.834522137 | 0.404 |
| hsa-mir-7976 | 1.345429717 | 1.61339 | 0.833915221 | 0.4043 |
| hsa-mir-1270 | 0.89190461 | 1.08908 | 0.818951324 | 0.4128 |
| hsa-mir-106b-3p | -0.24391259 | 0.30189 | -0.807948878 | 0.4191 |
| hsa-mir-485-3p | 0.960213735 | 1.19613 | 0.802767715 | 0.4221 |
| hsa-mir-487a-3p | 2.404471201 | 3.00377 | 0.800484584 | 0.4234 |
| hsa-mir-1180-3p | -0.603673575 | 0.76323 | -0.790944611 | 0.429 |
| hsa-mir-345-5p | -0.618622023 | 0.78307 | -0.789995676 | 0.4295 |
| hsa-mir-516a-1-5p | 2.346562509 | 3.00486 | 0.78092162 | 0.4348 |
| hsa-mir-516a-2-5p | 2.346562509 | 3.00486 | 0.78092162 | 0.4348 |
| hsa-mir-423-5p | 0.265452449 | 0.34008 | 0.780554926 | 0.4351 |
| hsa-mir-654-3p | 0.517190245 | 0.66347 | 0.779520054 | 0.4357 |
| hsa-mir-7977 | -2.323465293 | 2.98676 | -0.777920908 | 0.4366 |
| hsa-mir-146a-5p | 0.292640558 | 0.37764 | 0.774918786 | 0.4384 |
| hsa-mir-625-3p | -0.614544092 | 0.79599 | -0.772048374 | 0.4401 |
| hsa-mir-4661-5p | -2.298807596 | 2.98728 | -0.769532633 | 0.4416 |
| hsa-mir-483-3p | -2.298807596 | 2.98728 | -0.769532633 | 0.4416 |
| hsa-mir-29c-5p | -1.414377183 | 1.85754 | -0.761426398 | 0.4464 |
| hsa-mir-4326 | -2.235956155 | 2.98863 | -0.748154401 | 0.4544 |
| hsa-mir-185-5p | -0.230168037 | 0.30844 | -0.746243938 | 0.4555 |
| hsa-mir-126-5p | -0.243199403 | 0.32662 | -0.744602978 | 0.4565 |
| hsa-mir-769-5p | -0.509823028 | 0.69264 | -0.736058567 | 0.4617 |
| hsa-mir-3681-5p | -2.193899418 | 2.98957 | -0.733852009 | 0.463 |
| hsa-mir-190a-5p | -0.352988191 | 0.48208 | -0.732224631 | 0.464 |
| hsa-mir-331-3p | 1.051371835 | 1.43746 | 0.73141018 | 0.4645 |
| hsa-mir-655-3p | -2.173515652 | 2.99003 | -0.726920888 | 0.4673 |
| hsa-mir-365a-3p | -1.306469462 | 1.80288 | -0.724658675 | 0.4687 |
| hsa-mir-365b-3p | -1.306469462 | 1.80288 | -0.724658675 | 0.4687 |
| hsa-mir-4446-3p | 1.123634877 | 1.55339 | 0.723345334 | 0.4695 |
| hsa-mir-93-3p | 0.5497286 | 0.76406 | 0.719487353 | 0.4718 |
| hsa-mir-154-5p | -1.02578571 | 1.42638 | -0.719152417 | 0.472 |
| hsa-mir-199b-5p | 1.21151662 | 1.70902 | 0.708893628 | 0.4784 |
| hsa-mir-873-5p | -2.097533195 | 2.96756 | -0.706819739 | 0.4797 |
| hsa-mir-99b-5p | 0.246843379 | 0.34967 | 0.705937336 | 0.4802 |
| hsa-mir-17-3p | -1.232787866 | 1.75814 | -0.7011893 | 0.4832 |
| hsa-mir-4678 | -2.083543348 | 2.99216 | -0.696334902 | 0.4862 |
| hsa-mir-210-3p | 0.865804118 | 1.25034 | 0.692457356 | 0.4887 |
| hsa-mir-4742-5p | -2.071435596 | 2.99245 | -0.692219883 | 0.4888 |
| hsa-mir-34a-5p | -0.64929074 | 0.93959 | -0.691039862 | 0.4895 |
| hsa-mir-320c-1 | 0.426691848 | 0.6187 | 0.689659109 | 0.4904 |
| hsa-mir-320c-2 | 0.426691848 | 0.6187 | 0.689659109 | 0.4904 |
| hsa-mir-548j-3p | 2.067843758 | 3.00938 | 0.68713178 | 0.492 |
| hsa-mir-6758-5p | -2.036290668 | 2.99333 | -0.680276751 | 0.4963 |
| hsa-mir-10b-5p | -0.284500404 | 0.41887 | -0.679207531 | 0.497 |
| hsa-mir-181c-5p | -0.945489968 | 1.39344 | -0.678529961 | 0.4974 |
| hsa-mir-152-3p | 0.439566519 | 0.6494 | 0.676884471 | 0.4985 |
| hsa-mir-3127-3p | -2.02151234 | 2.9937 | -0.675255373 | 0.4995 |
| hsa-mir-5009-5p | -2.02151234 | 2.9937 | -0.675255373 | 0.4995 |
| hsa-mir-155-5p | -0.355229215 | 0.52646 | -0.674756468 | 0.4998 |
| hsa-mir-6503-5p | 2.016471232 | 3.0103 | 0.669856871 | 0.5029 |
| hsa-mir-1273h-3p | 0.488230451 | 0.738 | 0.661554526 | 0.5083 |
| hsa-mir-30d-5p | 0.126095871 | 0.1911 | 0.659850732 | 0.5093 |
| hsa-mir-501-3p | 0.29483556 | 0.44858 | 0.657260419 | 0.511 |
| hsa-mir-98-5p | -0.340355867 | 0.51904 | -0.655746369 | 0.512 |
| hsa-mir-184 | 1.046798261 | 1.60452 | 0.652404597 | 0.5141 |
| hsa-mir-136-3p | -0.913442197 | 1.40313 | -0.651004939 | 0.515 |
| hsa-mir-598-3p | 0.386072442 | 0.59872 | 0.644834766 | 0.519 |
| hsa-mir-671-5p | -0.483400253 | 0.7523 | -0.642565823 | 0.5205 |
| hsa-mir-4742-3p | -1.883028381 | 2.95101 | -0.638095667 | 0.5234 |
| hsa-mir-503-5p | -1.010920657 | 1.60432 | -0.630125956 | 0.5286 |
| hsa-mir-590-3p | 0.666525031 | 1.0589 | 0.629451142 | 0.5291 |
| hsa-mir-21-3p | -1.343827722 | 2.14787 | -0.625656698 | 0.5315 |
| hsa-mir-342-3p | 0.192250854 | 0.31149 | 0.61719589 | 0.5371 |
| hsa-mir-126-3p | 0.145621426 | 0.23645 | 0.615860607 | 0.538 |
| hsa-mir-205-5p | 0.485862517 | 0.79004 | 0.614981276 | 0.5386 |
| hsa-mir-652-3p | -0.412189548 | 0.67076 | -0.614514755 | 0.5389 |
| hsa-mir-22-3p | -0.161893383 | 0.26391 | -0.613435696 | 0.5396 |
| hsa-mir-589-5p | -0.402217999 | 0.65774 | -0.611516078 | 0.5409 |
| hsa-mir-23b-3p | 0.192870153 | 0.32039 | 0.601980524 | 0.5472 |
| hsa-mir-27a-5p | 0.418094296 | 0.69747 | 0.599446085 | 0.5489 |
| hsa-mir-369-5p | 0.609073747 | 1.02336 | 0.595171205 | 0.5517 |
| hsa-mir-181b-1-5p | -0.389720091 | 0.65799 | -0.592288056 | 0.5537 |
| hsa-mir-329-1-3p | -0.739377762 | 1.27165 | -0.581432077 | 0.5609 |
| hsa-mir-329-2-3p | -0.739377762 | 1.27165 | -0.581432077 | 0.5609 |
| hsa-mir-221-5p | -1.724260885 | 2.96776 | -0.580998008 | 0.5612 |
| hsa-mir-148b-3p | 0.173336744 | 0.30259 | 0.572845561 | 0.5667 |
| hsa-let-7i-5p | -0.143058833 | 0.25126 | -0.569357629 | 0.5691 |
| hsa-mir-1306-5p | -0.79813189 | 1.40199 | -0.569285436 | 0.5692 |
| hsa-mir-664a-3p | 1.087808509 | 1.91699 | 0.567455538 | 0.5704 |
| hsa-mir-106b-5p | 1.499065597 | 2.66243 | 0.563044657 | 0.5734 |
| hsa-mir-196a-1-5p | 1.408309876 | 2.53403 | 0.555758981 | 0.5784 |
| hsa-mir-27b-3p | -0.195802202 | 0.35482 | -0.55183852 | 0.5811 |
| hsa-mir-1250-5p | -1.646299192 | 2.98377 | -0.5517521 | 0.5811 |
| hsa-mir-642a-3p | 1.652291409 | 3.00404 | 0.55002376 | 0.5823 |
| hsa-mir-486-1-3p | 0.306589965 | 0.56971 | 0.538153083 | 0.5905 |
| hsa-mir-486-2-3p | 0.306589965 | 0.56971 | 0.538153083 | 0.5905 |
| hsa-mir-411-5p | 0.706681416 | 1.33458 | 0.529516604 | 0.5964 |
| hsa-mir-424-3p | 1.54737388 | 2.99936 | 0.515902149 | 0.6059 |
| hsa-mir-194-2-5p | -0.333446997 | 0.64684 | -0.515505336 | 0.6062 |
| hsa-mir-181a-1-3p | 0.38633716 | 0.75139 | 0.514164653 | 0.6071 |
| hsa-mir-26a-1-5p | -0.140095512 | 0.27326 | -0.512676456 | 0.6082 |
| hsa-mir-26a-2-5p | -0.140095512 | 0.27326 | -0.512676456 | 0.6082 |
| hsa-mir-192-5p | -0.207821705 | 0.41295 | -0.503256759 | 0.6148 |
| hsa-mir-379-5p | -0.264283003 | 0.52524 | -0.503162943 | 0.6148 |
| hsa-mir-3130-1-3p | 0.761605981 | 1.52356 | 0.49988525 | 0.6172 |
| hsa-mir-3130-2-3p | 0.761605981 | 1.52356 | 0.49988525 | 0.6172 |
| hsa-mir-431-5p | -0.315319536 | 0.63121 | -0.499545504 | 0.6174 |
| hsa-mir-382-5p | -0.268838559 | 0.54371 | -0.494453534 | 0.621 |
| hsa-mir-218-1-5p | -0.812220469 | 1.64653 | -0.493293533 | 0.6218 |
| hsa-mir-378d-1 | -0.731975264 | 1.49811 | -0.488599454 | 0.6251 |
| hsa-mir-378d-2 | -0.731975264 | 1.49811 | -0.488599454 | 0.6251 |
| hsa-mir-9-1-5p | 0.357021364 | 0.73602 | 0.485069267 | 0.6276 |
| hsa-mir-9-2-5p | 0.357021364 | 0.73602 | 0.485069267 | 0.6276 |
| hsa-mir-9-3-5p | 0.357021364 | 0.73602 | 0.485069267 | 0.6276 |
| hsa-mir-454-3p | -0.22147426 | 0.45739 | -0.484211756 | 0.6282 |
| hsa-mir-339-3p | 0.292705242 | 0.60595 | 0.483052342 | 0.6291 |
| hsa-mir-124-1-5p | -1.410974375 | 3.00191 | -0.470025274 | 0.6383 |
| hsa-mir-124-2-5p | -1.410974375 | 3.00191 | -0.470025274 | 0.6383 |
| hsa-mir-124-3-5p | -1.410974375 | 3.00191 | -0.470025274 | 0.6383 |
| hsa-mir-139-5p | -0.268734967 | 0.57226 | -0.469603394 | 0.6386 |
| hsa-mir-194-1-5p | -0.30229033 | 0.64538 | -0.468388695 | 0.6395 |
| hsa-mir-4665-5p | -1.389997878 | 2.97746 | -0.466839797 | 0.6406 |
| hsa-mir-708-5p | 0.861283474 | 1.84652 | 0.466435069 | 0.6409 |
| hsa-mir-218-2-5p | -0.765308463 | 1.64318 | -0.465749458 | 0.6414 |
| hsa-mir-1260b | -0.946859248 | 2.04054 | -0.464024449 | 0.6426 |
| hsa-mir-425-5p | 0.10800231 | 0.23479 | 0.459997189 | 0.6455 |
| hsa-mir-323a-3p | -0.801786885 | 1.77038 | -0.45289004 | 0.6506 |
| hsa-mir-3176 | -1.354686904 | 3.01108 | -0.449901175 | 0.6528 |
| hsa-mir-628-3p | 0.631970062 | 1.42557 | 0.44330956 | 0.6575 |
| hsa-mir-628-5p | -0.484288763 | 1.09945 | -0.440482523 | 0.6596 |
| hsa-mir-33a-5p | -1.238570987 | 2.81385 | -0.440169221 | 0.6598 |
| hsa-mir-9-1-3p | 0.398150802 | 0.908 | 0.438493396 | 0.661 |
| hsa-mir-9-2-3p | 0.398150802 | 0.908 | 0.438493396 | 0.661 |
| hsa-mir-9-3-3p | 0.398150802 | 0.908 | 0.438493396 | 0.661 |
| hsa-mir-296-5p | 0.923528743 | 2.11213 | 0.437250535 | 0.6619 |
| hsa-mir-877-5p | 1.273536945 | 2.94289 | 0.432750631 | 0.6652 |
| hsa-mir-92b-3p | -0.855936 | 1.98169 | -0.431921362 | 0.6658 |
| hsa-mir-92b-5p | 1.193293258 | 2.83568 | 0.420814447 | 0.6739 |
| hsa-mir-29b-1-3p | -0.167577243 | 0.40243 | -0.41641227 | 0.6771 |
| hsa-mir-29b-2-3p | -0.167577243 | 0.40243 | -0.41641227 | 0.6771 |
| hsa-mir-450b-5p | -1.236859535 | 2.97925 | -0.415157401 | 0.678 |
| hsa-mir-378a-5p | 0.590634827 | 1.4317 | 0.412540322 | 0.6799 |
| hsa-mir-1185-1-5p | 1.194490473 | 2.96794 | 0.402464784 | 0.6873 |
| hsa-mir-1185-2-5p | 1.194490473 | 2.96794 | 0.402464784 | 0.6873 |
| hsa-mir-454-5p | 0.535702662 | 1.3531 | 0.39590723 | 0.6922 |
| hsa-mir-487b-3p | 0.37265413 | 0.94129 | 0.395899116 | 0.6922 |
| hsa-mir-4723-5p | -1.188198733 | 3.0152 | -0.394070179 | 0.6935 |
| hsa-mir-204-5p | -0.535268441 | 1.37993 | -0.387895053 | 0.6981 |
| hsa-mir-150-5p | -0.150657505 | 0.39161 | -0.384717218 | 0.7004 |
| hsa-mir-1249-3p | 1.133195781 | 2.99028 | 0.378959203 | 0.7047 |
| hsa-mir-548k | -0.993771957 | 2.66054 | -0.373522078 | 0.7088 |
| hsa-mir-338-3p | 0.226527881 | 0.60706 | 0.373156841 | 0.709 |
| hsa-mir-1271-5p | -0.488341498 | 1.31853 | -0.370367992 | 0.7111 |
| hsa-let-7e-5p | 0.143747211 | 0.38982 | 0.368751661 | 0.7123 |
| hsa-mir-3920 | -1.101310608 | 3.01714 | -0.365018635 | 0.7151 |
| hsa-mir-548e-5p | -0.819334678 | 2.25696 | -0.363026007 | 0.7166 |
| hsa-mir-941-1 | -0.182209465 | 0.50333 | -0.36201006 | 0.7173 |
| hsa-mir-941-2 | -0.182209465 | 0.50333 | -0.36201006 | 0.7173 |
| hsa-mir-409-5p | 0.680943702 | 1.8978 | 0.358807202 | 0.7197 |
| hsa-mir-3614-5p | 0.634663711 | 1.78804 | 0.354949407 | 0.7226 |
| hsa-mir-493-5p | 0.450477454 | 1.29233 | 0.348578557 | 0.7274 |
| hsa-mir-99a-5p | 0.161244153 | 0.46346 | 0.347914996 | 0.7279 |
| hsa-mir-135a-1-5p | -0.626130876 | 1.80199 | -0.347467306 | 0.7282 |
| hsa-mir-135a-2-5p | -0.626130876 | 1.80199 | -0.347467306 | 0.7282 |
| hsa-mir-330-5p | -0.51422637 | 1.53126 | -0.335818277 | 0.737 |
| hsa-mir-574-3p | 0.232168375 | 0.69746 | 0.332877743 | 0.7392 |
| hsa-mir-409-3p | -0.152042489 | 0.47164 | -0.322369532 | 0.7472 |
| hsa-mir-125a-5p | -0.141179584 | 0.44694 | -0.315880942 | 0.7521 |
| hsa-mir-2110 | 0.25926009 | 0.82784 | 0.31317781 | 0.7541 |
| hsa-mir-3688-1-3p | -0.75527507 | 2.43184 | -0.310577523 | 0.7561 |
| hsa-mir-3688-2-3p | -0.75527507 | 2.43184 | -0.310577523 | 0.7561 |
| hsa-mir-550a-1-3p | -0.602502493 | 1.99701 | -0.301702576 | 0.7629 |
| hsa-mir-550a-2-3p | -0.602502493 | 1.99701 | -0.301702576 | 0.7629 |
| hsa-mir-550a-3-3p | -0.602502493 | 1.99701 | -0.301702576 | 0.7629 |
| hsa-mir-197-3p | 0.153398128 | 0.51122 | 0.300061307 | 0.7641 |
| hsa-mir-181b-2-5p | -0.182488695 | 0.60921 | -0.29954761 | 0.7645 |
| hsa-mir-4524a-3p | -0.878056149 | 2.95944 | -0.296696678 | 0.7667 |
| hsa-mir-889-3p | 0.540575376 | 1.83882 | 0.293978754 | 0.7688 |
| hsa-mir-148a-3p | -0.088458099 | 0.30816 | -0.28705654 | 0.7741 |
| hsa-mir-30d-3p | -0.623222512 | 2.19074 | -0.284480133 | 0.776 |
| hsa-mir-382-3p | 0.558149491 | 1.96525 | 0.284009175 | 0.7764 |
| hsa-mir-377-3p | -0.731130761 | 2.59242 | -0.282026864 | 0.7779 |
| hsa-mir-30a-3p | 0.350269389 | 1.25738 | 0.278570012 | 0.7806 |
| hsa-mir-335-5p | 0.135560436 | 0.48855 | 0.277476201 | 0.7814 |
| hsa-mir-125b-2-5p | 0.137698835 | 0.49683 | 0.277153886 | 0.7817 |
| hsa-mir-138-2-5p | -0.787970244 | 2.84755 | -0.276718912 | 0.782 |
| hsa-mir-141-3p | -0.23308995 | 0.84817 | -0.274813807 | 0.7835 |
| hsa-mir-6721-5p | -0.767800046 | 2.82084 | -0.272188286 | 0.7855 |
| hsa-mir-195-5p | -0.25774946 | 0.95325 | -0.270390027 | 0.7869 |
| hsa-mir-374a-5p | 0.170063489 | 0.63203 | 0.269073312 | 0.7879 |
| hsa-mir-197-5p | -0.542442856 | 2.07823 | -0.261012213 | 0.7941 |
| hsa-mir-3124-5p | 0.76407435 | 2.94774 | 0.259206695 | 0.7955 |
| hsa-mir-146b-5p | 0.092016672 | 0.35672 | 0.257949713 | 0.7964 |
| hsa-mir-3679-5p | -0.625557179 | 2.43197 | -0.257222016 | 0.797 |
| hsa-mir-874-5p | 0.616859253 | 2.41218 | 0.255726693 | 0.7982 |
| hsa-mir-1179 | -0.49135462 | 1.94469 | -0.252664165 | 0.8005 |
| hsa-mir-124-1-3p | 0.482188186 | 1.92368 | 0.250659414 | 0.8021 |
| hsa-mir-124-2-3p | 0.482188186 | 1.92368 | 0.250659414 | 0.8021 |
| hsa-mir-124-3-3p | 0.482188186 | 1.92368 | 0.250659414 | 0.8021 |
| hsa-mir-130a-3p | -0.140026631 | 0.56045 | -0.24984708 | 0.8027 |
| hsa-mir-502-3p | -0.143480548 | 0.57639 | -0.248931044 | 0.8034 |
| hsa-mir-125b-1-5p | 0.120015343 | 0.49182 | 0.244022767 | 0.8072 |
| hsa-mir-196a-2-5p | -0.507267291 | 2.0795 | -0.243937598 | 0.8073 |
| hsa-mir-219a-2-5p | 0.712629642 | 2.99412 | 0.238009665 | 0.8119 |
| hsa-mir-671-3p | 0.138520892 | 0.59965 | 0.231002129 | 0.8173 |
| hsa-mir-320d-2 | -0.158147063 | 0.68634 | -0.230422028 | 0.8178 |
| hsa-mir-337-5p | 0.239735765 | 1.04949 | 0.228430508 | 0.8193 |
| hsa-mir-1908-5p | 0.487563816 | 2.13882 | 0.227959557 | 0.8197 |
| hsa-mir-103a-1-3p | -0.047995252 | 0.21299 | -0.225338028 | 0.8217 |
| hsa-mir-103a-2-3p | -0.047995252 | 0.21299 | -0.225338028 | 0.8217 |
| hsa-mir-501-5p | -0.535132485 | 2.37864 | -0.22497376 | 0.822 |
| hsa-mir-191-3p | 0.656897604 | 2.94962 | 0.222705971 | 0.8238 |
| hsa-mir-17-5p | 0.081462071 | 0.36693 | 0.22201034 | 0.8243 |
| hsa-mir-361-5p | 0.110486771 | 0.50887 | 0.217122356 | 0.8281 |
| hsa-mir-145-5p | 0.131275831 | 0.61236 | 0.214375798 | 0.8303 |
| hsa-mir-942-5p | -0.186949401 | 0.87306 | -0.214131821 | 0.8304 |
| hsa-mir-3913-1-5p | 0.275335027 | 1.31443 | 0.209470916 | 0.8341 |
| hsa-mir-3913-2-5p | 0.275335027 | 1.31443 | 0.209470916 | 0.8341 |
| hsa-mir-425-3p | 0.208508641 | 1.00766 | 0.206923005 | 0.8361 |
| hsa-mir-219a-1-5p | -0.457785372 | 2.23384 | -0.204932418 | 0.8376 |
| hsa-mir-421 | 0.178352646 | 0.88011 | 0.202648718 | 0.8394 |
| hsa-mir-625-5p | 0.165197769 | 0.82364 | 0.200571072 | 0.841 |
| hsa-mir-766-3p | 0.161657598 | 0.8352 | 0.193555395 | 0.8465 |
| hsa-mir-30c-1-5p | 0.050705336 | 0.26416 | 0.191948431 | 0.8478 |
| hsa-mir-30c-2-5p | 0.050705336 | 0.26416 | 0.191948431 | 0.8478 |
| hsa-mir-153-1-3p | 0.475025002 | 2.49176 | 0.190638363 | 0.8488 |
| hsa-mir-153-2-3p | 0.475025002 | 2.49176 | 0.190638363 | 0.8488 |
| hsa-mir-22-5p | -0.329576949 | 1.73193 | -0.190294096 | 0.8491 |
| hsa-mir-7706 | 0.445336595 | 2.34419 | 0.189974285 | 0.8493 |
| hsa-mir-432-5p | 0.0853604 | 0.46819 | 0.182319081 | 0.8553 |
| hsa-mir-323b-3p | -0.124975916 | 0.69286 | -0.180376258 | 0.8569 |
| hsa-mir-1292-5p | 0.324230503 | 1.80178 | 0.179949693 | 0.8572 |
| hsa-mir-18a-3p | -0.209754771 | 1.2203 | -0.171887362 | 0.8635 |
| hsa-mir-149-5p | 0.331182193 | 1.92726 | 0.17184062 | 0.8636 |
| hsa-mir-769-3p | 0.272051837 | 1.61029 | 0.168945583 | 0.8658 |
| hsa-mir-374a-3p | -0.235114845 | 1.47087 | -0.15984794 | 0.873 |
| hsa-mir-488-3p | -0.450880825 | 2.83283 | -0.159162599 | 0.8735 |
| hsa-mir-4532 | 0.17537398 | 1.11159 | 0.157768925 | 0.8746 |
| hsa-mir-193a-5p | -0.164043026 | 1.06618 | -0.153860555 | 0.8777 |
| hsa-mir-20a-5p | -0.053223071 | 0.37741 | -0.14102141 | 0.8879 |
| hsa-mir-1343-3p | 0.360156224 | 2.64176 | 0.136331803 | 0.8916 |
| hsa-mir-6842-3p | -0.242853852 | 1.94235 | -0.125030626 | 0.9005 |
| hsa-mir-548j-5p | 0.061951991 | 0.53279 | 0.116279214 | 0.9074 |
| hsa-mir-151b | -0.151239025 | 1.30267 | -0.116099687 | 0.9076 |
| hsa-mir-137 | 0.145735852 | 1.28179 | 0.113696964 | 0.9095 |
| hsa-mir-186-5p | -0.031917712 | 0.28284 | -0.112848629 | 0.9102 |
| hsa-mir-381-3p | 0.075254128 | 0.6834 | 0.11011795 | 0.9123 |
| hsa-mir-4286 | 0.276581114 | 2.51994 | 0.109757068 | 0.9126 |
| hsa-mir-664a-5p | -0.058254704 | 0.57138 | -0.101955132 | 0.9188 |
| hsa-mir-494-3p | 0.120522326 | 1.22566 | 0.098332896 | 0.9217 |
| hsa-mir-3074-5p | -0.283682045 | 2.9451 | -0.096323523 | 0.9233 |
| hsa-mir-340-3p | -0.269449636 | 2.95168 | -0.091286846 | 0.9273 |
| hsa-mir-874-3p | -0.094729951 | 1.04442 | -0.090701114 | 0.9277 |
| hsa-mir-138-1-5p | 0.132987358 | 1.53486 | 0.086644731 | 0.931 |
| hsa-mir-127-3p | 0.047398045 | 0.56541 | 0.08382895 | 0.9332 |
| hsa-mir-320d-1 | -0.055253658 | 0.70456 | -0.078422617 | 0.9375 |
| hsa-mir-4772-3p | 0.181243176 | 2.95363 | 0.061362949 | 0.9511 |
| hsa-mir-4677-3p | 0.104509155 | 1.74255 | 0.059974953 | 0.9522 |
| hsa-let-7i-3p | -0.136698022 | 2.41953 | -0.056497742 | 0.9549 |
| hsa-mir-1224-5p | 0.098866459 | 1.8148 | 0.054477913 | 0.9566 |
| hsa-mir-369-3p | -0.099758769 | 1.9193 | -0.051976646 | 0.9585 |
| hsa-mir-100-5p | 0.038706289 | 0.78553 | 0.049274107 | 0.9607 |
| hsa-mir-25-5p | -0.052735013 | 1.09626 | -0.048104398 | 0.9616 |
| hsa-mir-532-3p | -0.045729343 | 0.96097 | -0.047586466 | 0.962 |
| hsa-mir-485-5p | 0.060698119 | 1.3408 | 0.045269914 | 0.9639 |
| hsa-mir-6852-5p | 0.051623902 | 1.33638 | 0.038629677 | 0.9692 |
| hsa-mir-362-3p | -0.089711666 | 2.33758 | -0.038378036 | 0.9694 |
| hsa-mir-132-3p | 0.055712767 | 1.45477 | 0.038296696 | 0.9695 |
| hsa-mir-493-3p | 0.037401333 | 0.97916 | 0.038197267 | 0.9695 |
| hsa-mir-3615 | -0.015525649 | 0.45228 | -0.03432759 | 0.9726 |
| hsa-mir-223-5p | 0.055189522 | 1.76859 | 0.031205293 | 0.9751 |
| hsa-mir-200a-3p | 0.065004144 | 2.14037 | 0.030370548 | 0.9758 |
| hsa-mir-200c-3p | 0.029119794 | 1.04211 | 0.027943242 | 0.9777 |
| hsa-mir-548o-2-3p | -0.076854899 | 2.93694 | -0.02616832 | 0.9791 |
| hsa-mir-548o-3p | -0.076854899 | 2.93694 | -0.02616832 | 0.9791 |
| hsa-mir-885-5p | -0.076264255 | 3.00319 | -0.025394391 | 0.9797 |
| hsa-mir-301a-3p | -0.019982366 | 0.81673 | -0.024466224 | 0.9805 |
| hsa-mir-129-1-5p | -0.065146793 | 2.90104 | -0.022456342 | 0.9821 |
| hsa-mir-129-2-5p | -0.065146793 | 2.90104 | -0.022456342 | 0.9821 |
| hsa-mir-505-3p | 0.019771952 | 1.02532 | 0.019283729 | 0.9846 |
| hsa-mir-95-3p | -0.015299354 | 0.83069 | -0.018417537 | 0.9853 |
| hsa-mir-134-5p | -0.011557542 | 0.64621 | -0.017885187 | 0.9857 |
| hsa-mir-370-3p | -0.010172104 | 0.62974 | -0.016152858 | 0.9871 |
| hsa-mir-548a-1-3p | 0.018906543 | 2.1648 | 0.008733612 | 0.993 |
| hsa-mir-548a-2-3p | 0.018906543 | 2.1648 | 0.008733612 | 0.993 |
| hsa-mir-548a-3-3p | 0.018906543 | 2.1648 | 0.008733612 | 0.993 |
| hsa-mir-376a-1-3p | 0.003327375 | 1.48629 | 0.00223871 | 0.9982 |
| hsa-mir-376a-2-3p | 0.003327375 | 1.48629 | 0.00223871 | 0.9982 |

**Supplementary Table 2:** Normalized exosomal miRNA expression differences between CLL-FIT and CLL-UNFIT

| miRNA | CLL-Fit | CLL-Unfit | Log2 Fold Difference (SE) | Wald Test Statistic | Wald Test *p*-value |
| --- | --- | --- | --- | --- | --- |
| hsa-mir-101-2-3p | 358.8 ± 110.7 | 709.1 ± 695.3 | -0.98 (0.37) | -2.68 | 0.0074 |
| hsa-mir-199a (1-3p and 2-3p) | 2000.1 ± 806.3 | 1086.1 ± 597.8 | 0.88 (0.33) | 2.67 | 0.0076 |
| hsa-mir-199b-3p | 2000.1 ± 806.3 | 1086.1 ± 597.8 | 0.88 (0.33) | 2.67 | 0.0076 |
| hsa-mir-101-1-3p | 358.8 ± 110.7 | 702.8 ± 699.8 | -0.97 (0.37) | -2.60 | 0.0093 |
| hsa-mir-378a-3p | 39.3 ± 31.4 | 192.0 ± 353.6 | -2.28 (0.90) | -2.54 | 0.0112 |
| hsa-mir-32-5p | 6.7 ± 8.0 | 38.1 ± 51.7 | -2.47 (0.98) | -2.51 | 0.0119 |
| hsa-mir-24 (1-3p and 2-3p) | 231.5 ± 67.5 | 137.2 ± 69.1 | 0.76 (0.31) | 2.48 | 0.013 |
| hsa-mir-130b-5p | 18.9 ± 16.0 | 2.9 ± 5.1 | 2.70 (1.09) | 2.47 | 0.0133 |
| hsa-mir-744-5p | 275.1 ± 148.2 | 141.2 ± 90.8 | 0.97 (0.39) | 2.46 | 0.0138 |
| hsa-mir-1301-3p | 23.2 ± 25.9 | 2.6 ± 5.6 | 3.15 (1.31) | 2.39 | 0.0167 |
| hsa-mir-328-3p | 69.0 ± 90.2 | 22.6 ± 20.4 | 1.61 (0.69) | 2.35 | 0.0189 |
| hsa-mir-4433b-3p | 19.3 ± 22.1 | 2.1 ± 3.7 | 3.14 (1.35) | 2.33 | 0.0199 |
| hsa-mir-4433b-5p | 109.4 ± 88.8 | 41.9 ± 30.5 | 1.39 (0.60) | 2.30 | 0.0212 |
| hsa-mir-383-5p | 8.2 ± 24.7 | 0.0 ± 0.0 | 5.67 (2.47) | 2.29 | 0.022 |
| hsa-mir-29c-3p | 895.7 ± 464.6 | 1879.8 ± 2799.6 | -1.07 (0.48) | -2.25 | 0.0247 |
| hsa-mir-16 (1-5p and 2-5p) | 24287.9 ± 14781.3 | 40209.4 ± 18457.5 | -0.73 (0.34) | -2.16 | 0.0304 |
| hsa-mir-151a-5p | 70.8 ± 39.3 | 35.6 ± 30.5 | 0.98 (0.45) | 2.15 | 0.0313 |
| hsa-let-7f-1-5p | 3920.2 ± 1591.7 | 6090.8 ± 2622.7 | -0.64 (0.30) | -2.15 | 0.0317 |
| hsa-mir-19b (1-3p and 2-3p) | 143.2 ± 83.7 | 286.4 ± 318.1 | -1.00 (0.47) | -2.13 | 0.0329 |
| hsa-mir-183-5p | 76.8 ± 64.0 | 156.7 ± 136.5 | -1.02 (0.48) | -2.12 | 0.0337 |
| hsa-mir-451a | 4683.4 ± 2758.3 | 8650.8 ± 4511.6 | -0.89 (0.43) | -2.08 | 0.0375 |
| hsa-mir-324-3p | 7.7 ± 15.0 | 0.8 ± 1.2 | 3.35 (1.63) | 2.05 | 0.0404 |
| hsa-let-7f-2-5p | 4069.2 ± 1621.0 | 6127.0 ± 2571.2 | -0.59 (0.29) | -2.05 | 0.0407 |
| hsa-mir-151a-3p | 2250.9 ± 610.4 | 1503.6 ± 788.9 | 0.58 (0.29) | 2.04 | 0.0415 |
| hsa-mir-576-5p | 31.0 ± 23.4 | 65.8 ± 44.6 | -1.06 (0.53) | -2.02 | 0.0436 |
| hsa-mir-6772-3p | 10.0 ± 24.2 | 0.0 ± 0.0 | 5.97 (2.96) | 2.02 | 0.0437 |
| hsa-mir-182-5p | 366.6 ± 229.6 | 655.2 ± 506.6 | -0.84 (0.42) | -1.97 | 0.0486 |
| hsa-mir-1296-5p | 6.8 ± 9.1 | 0.4 ± 1.3 | 3.73 (1.89) | 1.97 | 0.0486 |

Data are mean ± SD of normalized expression values, log<sub>2</sub> fold change and SE for CLL-FIT/CLL-UNFIT.

**Supplementary Table 3.**
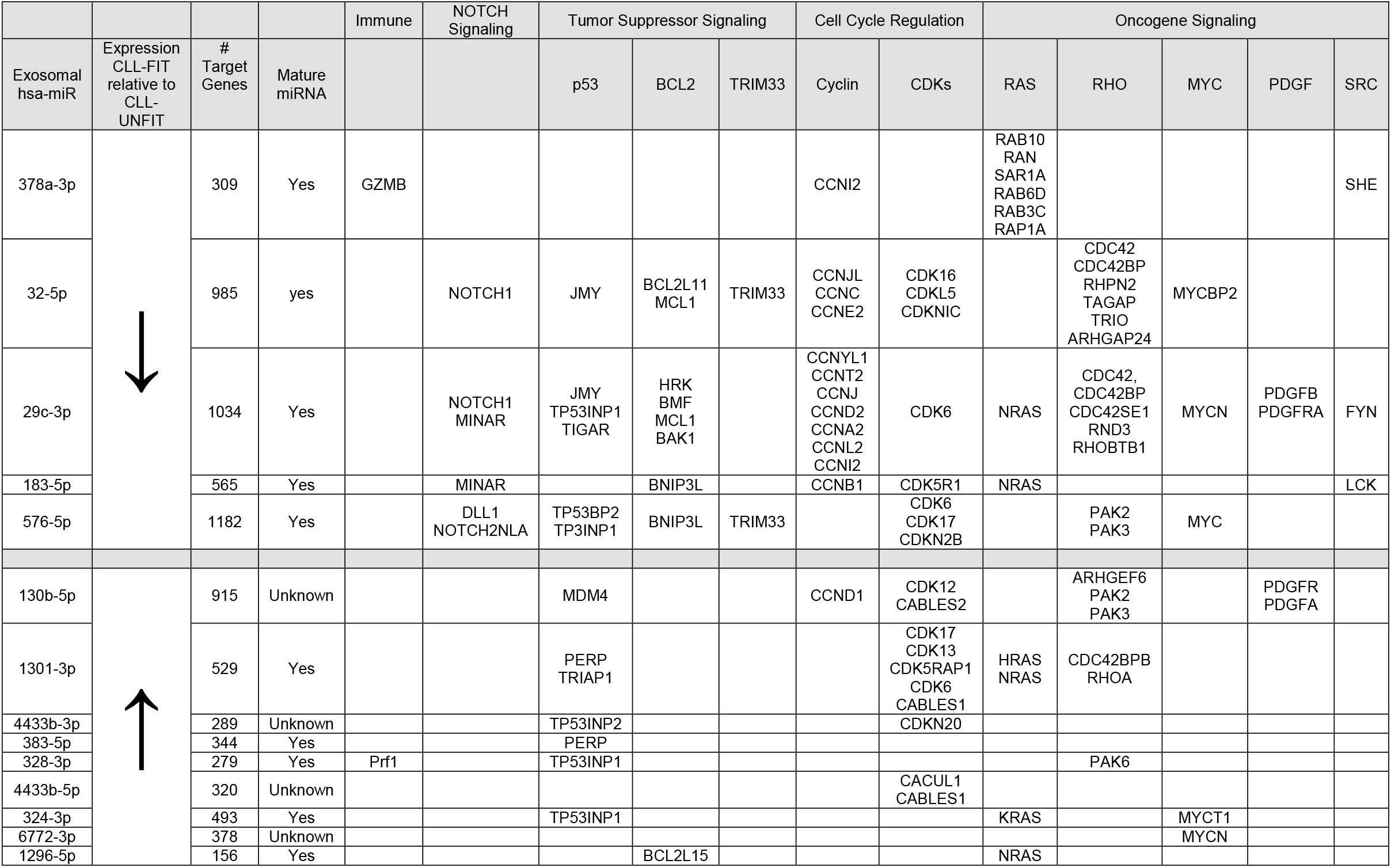
miRNA target genes associated with CLL. Acquired from miRDB (http://mirdb.org/index.html) accessed on December 1^st^ 2020

**Supplementary Table 4:**
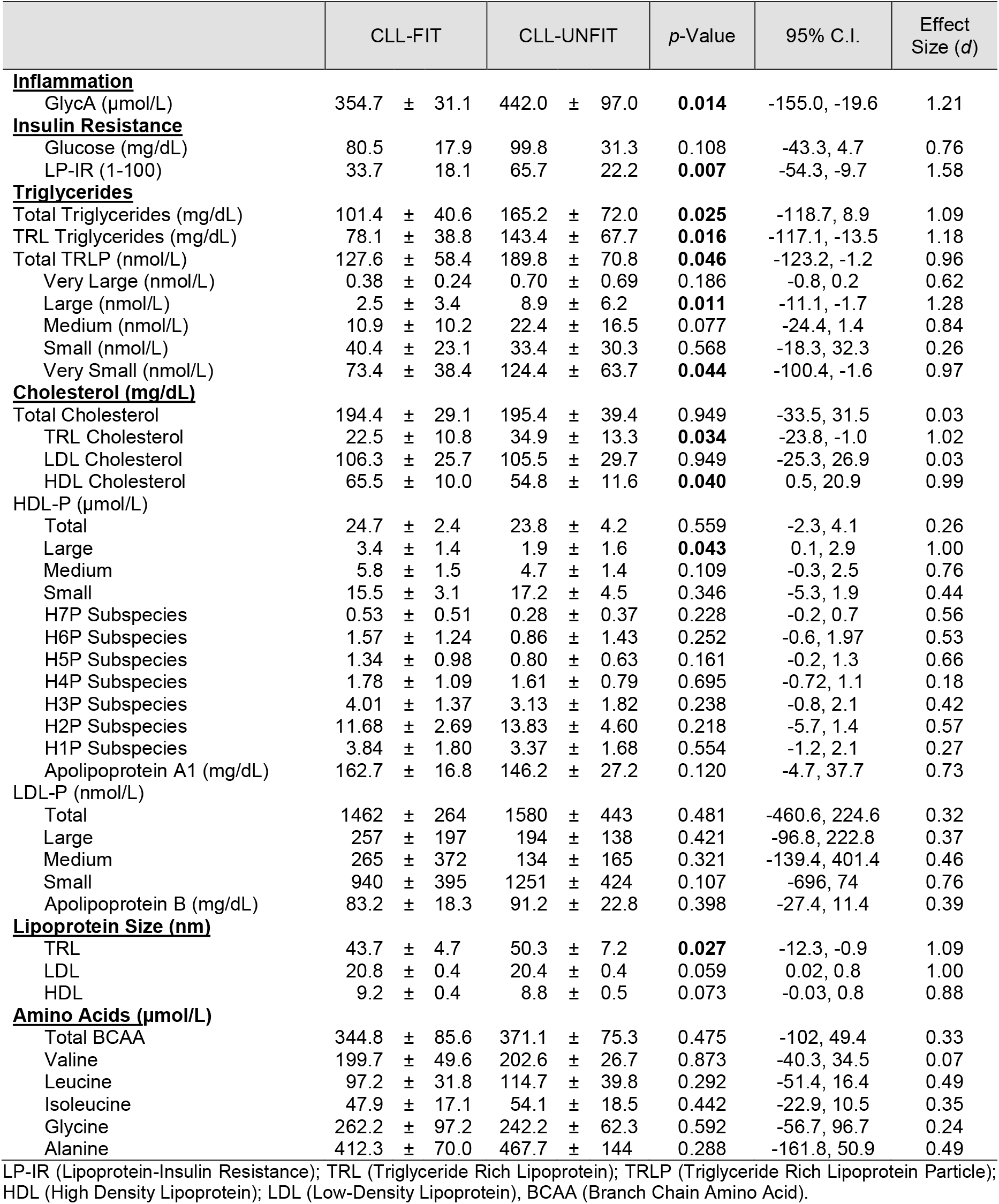
Complete plasma NMR measurements of systemic inflammation, insulin resistance, lipids, lipoproteins, and amino acids

